# Fairness and efficiency considerations in COVID-19 vaccine allocation strategies: a case study comparing front-line workers and 65-74 year olds in the United States

**DOI:** 10.1101/2022.02.03.22270414

**Authors:** Eva Rumpler, Justin M. Feldman, Mary T. Bassett, Marc Lipsitch

## Abstract

The COVID-19 epidemic in the United States has been characterized by two stark disparities. COVID-19 burden has been unequally distributed among racial and ethnic groups and at the same time the mortality rates have been sharply higher among older age groups. These disparities have led some to suggest that inequalities could be reduced by vaccinating front-line workers before vaccinating older individuals, as older individuals in the US are disproportionately Non-Hispanic White.

We compare the performance of two distribution policies, one allocating vaccines to front-line workers and another to older individuals aged 65-74-year-old. We estimate both the number of lives saved and the number of years of life saved under each of the policies, overall and in every race/ethnicity groups, in the United States and every state.

We show that prioritizing COVID-19 vaccines for 65-74-year-olds saves both more lives and more years of life than allocating vaccines front-line workers in each racial/ethnic group, in the United States as a whole and in nearly every state. When evaluating fairness of vaccine allocation policies, the overall benefit to impact of each population subgroup should be considered, not only the proportion of doses that is distributed to each subgroup. Further work can identify prioritization schemes that perform better on multiple equity metrics.

## 2 Introduction

Two stark disparities define the mortality impact of the COVID-19 epidemic in the United States. The risk of death given infection (infection fatality rate) rises sharply with age. By one estimate, this increase is exponential, about 10-fold for each 19-year increase in age [1]. Another estimate fits a linear increase and finds a 1 ·18 percentage-point absolute increase in infection fatality rate per decade of age [2]. At the same time, Black, Hispanic, and Native American persons in the US have experienced confirmed infections, hospitalizations, and deaths at dramatically higher rates than White or Asian persons [3], reflecting a pattern of racial/ethnic and socioeconomic disparity that is observed also in other populations [4–7]. As of March 2021, White and Asian populations in the US have similar age-adjusted COVID-19 mortality rates (respectively 121/100,000 and 117/100,000). Compared to White populations, the age-adjusted COVID-19 mortality rate is 1 ·7-fold higher among Black populations (241/100,000), 1 ·9-fold higher among Indigenous populations (263/100,000), 2 ·0-fold higher among Hispanic populations (287/100,000) and 2 ·2-fold higher among Pacific Islander populations (312 / 100,000) [8]. An additional relevant fact that connects these two disparities is that White and Asian populations are older on average than Black, Hispanic, and other racial/ethnic groups in the United States.

The two risk factors of age and race, as well as their interaction, have received attention in discussions on vaccine prioritization. While prioritization of older adults and individuals with comorbid conditions predisposing them to higher infection-fatality rates is under most scenarios expected to save more lives in the aggregate than non-prioritized strategies [9–12], such a strategy in the US involves providing more vaccine doses, earlier, to White individuals and fewer to Black, Hispanic, Asian and Native American people, due to the differences in age distribution. An alternative strategy, that of prioritizing essential or front-line workers, has been discussed as a way to provide more vaccine doses to those racial/ethnic groups that are hardest hit by COVID-19, because essential workers are more ethnically and racially diverse than older individuals in the US.

Here we quantify the potential impacts overall and by race/ethnicity, comparing two groups that have been widely discussed for prioritization below long-term care residents and health care workers and those 75 and older, but above other members of the population: i) individuals 65-74 not in long-term care or nursing homes, and ii) front-line (non health care) workers. For the United States and each state individually, we quantify the extent to which these two policies would provide vaccines to members of different racial and ethnic groups, and we ask the further question of how the two policies compare in terms of expected lives and years of life saved overall and for each racial/ethnic group.

Such estimates require certain modeling assumptions. We outline those in detail in Methods, but we emphasize here the most important. First, we consider only the direct effects of vaccination in preventing death from COVID-19 in vaccinated persons. Indirect effects on transmission are not considered in this study, to simplify and make transparent the analysis and because vaccines’ effects on transmission were not clear at the time of vaccine allocation decisions. Second, we make the assumption that from the moment the prioritization decision is made, future COVID-19 deaths in strata defined by age group, race/ethnicity and state, and occupation (essential worker or not) are proportional to those estimated for the period prior to January 30, 2021. This assumption links future disparities in death rates related to all these factors to past ones. Third, we assume that vaccine uptake and effectiveness is even across race/ethnic groups within an age group and state under each policy; we discuss the impact of this assumption later. The second and third assumptions together enable a comparison of the projected number of lives saved in each race/ethnicity, by state, by allocating a fixed number of vaccine doses under each of the two policies.

## 3 Methods

### 3.1 Notation

We repeat our analyses across multiple age categories, race/ethnicity groups and US states. We consider various geographical entities *k*: either the entirety of the United States, or any one of 50 states or the District of Columbia. The age groups considered are either 65-74 year old for policy *S*, or 16-64 (further detailed into 16-24, 25-34, 35-44, 45-54, 55-64) for front-line workers for policy *F*. We conducted our analyses for seven race/ethnicity categories (Non-Hispanic Whites, Hispanics, Non-Hispanic Blacks or African American, Non-Hispanic Asians, Non-Hispanic Two or more races, Non-Hispanic American Indian or Alaska Native, Non-Hispanic Native Hawaiian or other Pacific Islander).

Notations used are detailed in Table 1 below.

**Table 1:**
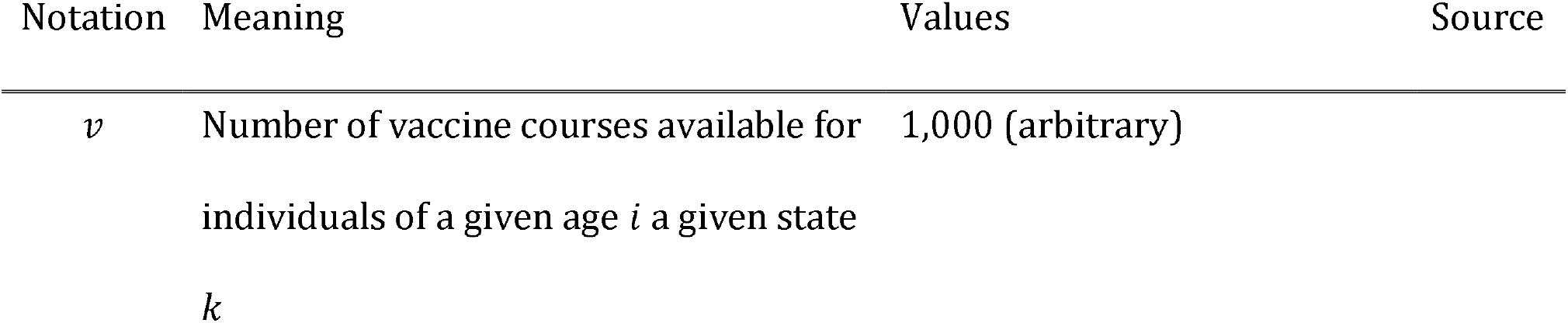
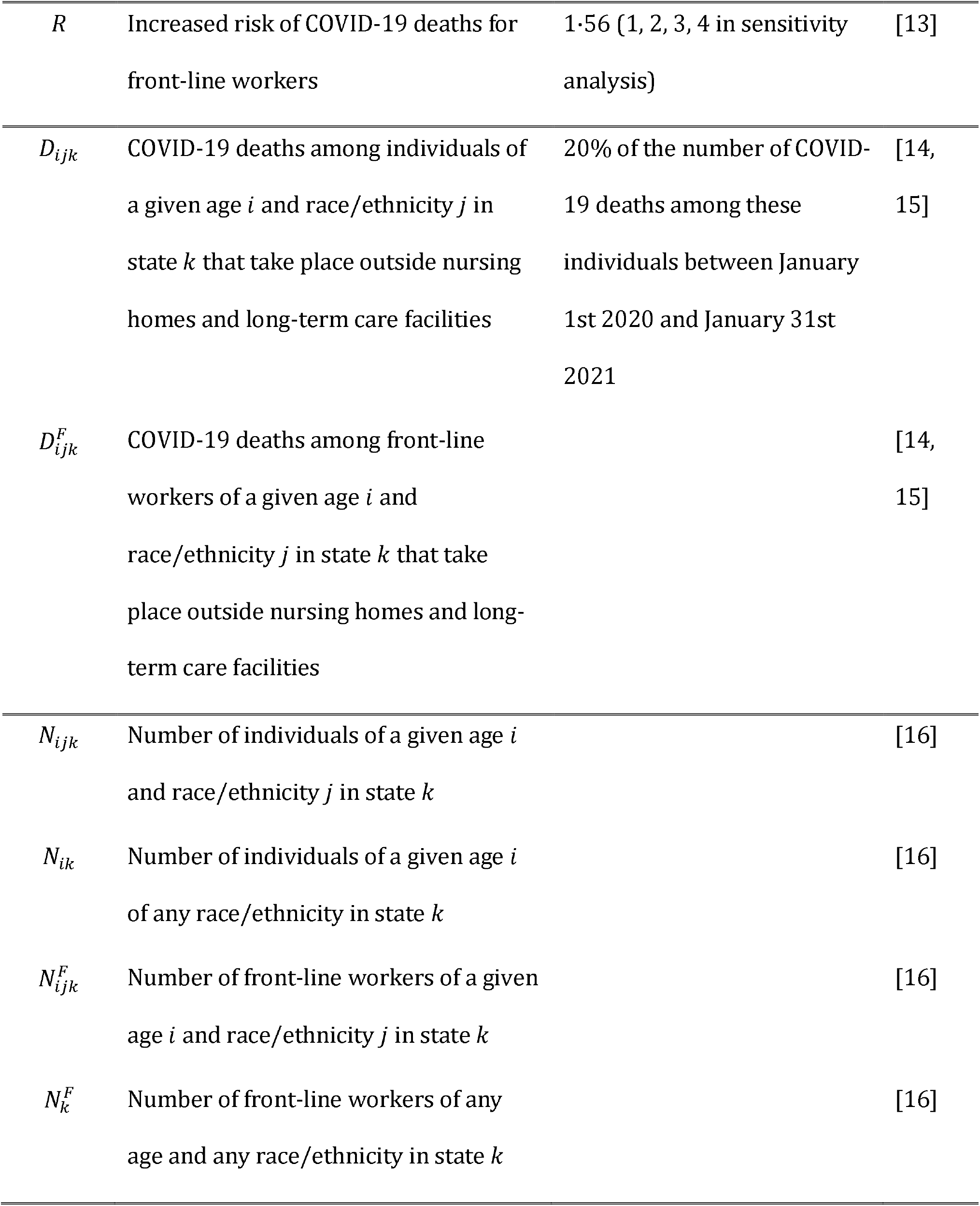
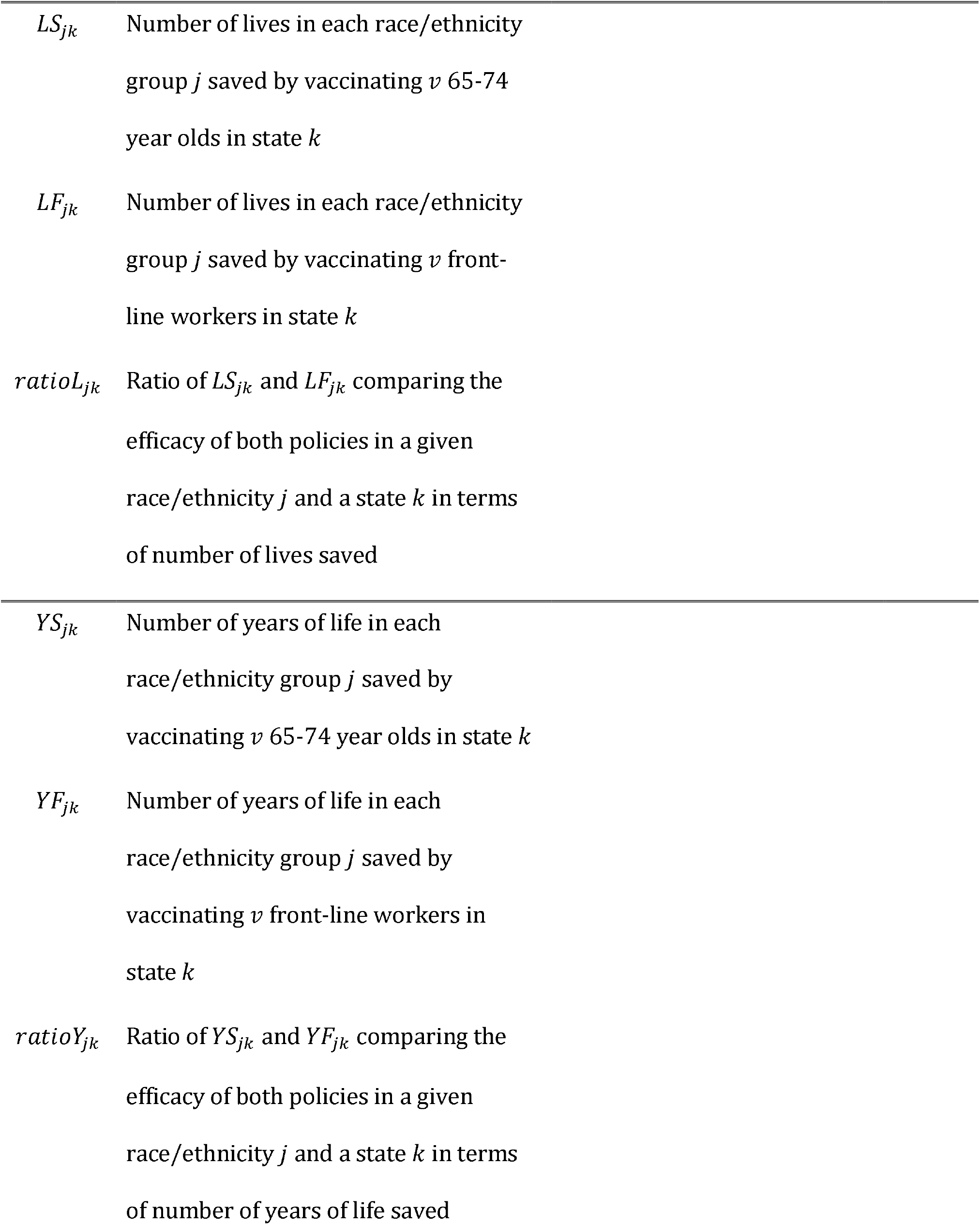
Notation and input values.

### 3.2 Data sources and estimation

#### 3.2.1 Estimating the number of COVID-19 deaths outside of nursing homes and long term care facilities by state, race/ethnicity and age categories

We obtained the number of COVID-19 deaths by state, age, race and Hispanic origin group reported to the National Center for Health Statistics (NCHS) between January 1st 2020 and January 30th 2021 [14]. We also extracted the proportion of COVID-19 deaths that took place outside of nursing homes and long term care facilities between January 4th 2020 and January 30th 2021, by state and age categories from the NCHS [15]. Deaths counts ranging between 1 and 9, which were not reported due to the NCHS confidentiality standards, were approximated by the median value 5.

As nursing home and long-term care facilities residents and employees have access to vaccination before 65-74 and front-line workers, we do not consider deaths that took place in those settings as part of our analysis. Assuming that this proportion of COVID-19 deaths in nursing homes and long term care facilities is constant across races and ethnicity for a given state and age category, we compute the proportion of COVID-19 deaths that took place outside of these settings for each joint stratum of state, age category and race.

#### 3.2.2 Extracting the number of front-line workers and total number of individuals by state, race and age categories

We obtained the number of individuals in each US state, race and age category from the American Community Survey (ACS) estimates for the year 2019 [16]. The demographic variables we included were State, Age, Occupation, Race and Hispanic origin. We categorized these into seven race/ethnicity groups: Non-Hispanic Whites, Hispanics, Non-Hispanic Blacks or African American, Non-Hispanic Asians, Non-Hispanic Two or more races, Non-Hispanic American Indian or Alaska Native, Non-Hispanic Native Hawaiian or other Pacific Islander.

For each state, race/ethnicity, and 10-year age category 16-64, we also construct the number of front-line workers employed in occupation categories corresponding to the definition of workers who are likely to be exposed to COVID-19 as defined by the state of Massachusetts [17]. This list includes K-12 educators, drivers, retail, funeral, food and beverage workers. Summary tables showing the occupations included, as well as the share of individuals from each race/ethnicity in each occupation can be found in S1 Table.

#### 3.2.3 Estimating the mortality rate among front-line workers

Because of higher exposure to infection and possibly other factors, front-line workers may experience a higher risk of COVID-19 death than other individuals of the same age, race/ethnicity, and state. To estimate the increased mortality rate among front-line workers, we multiply the mortality rate of a given age/state/race category by *R*, the increased risk due to their increased exposure to SARS-CoV-2 at work and possible increased mortality from other factors.

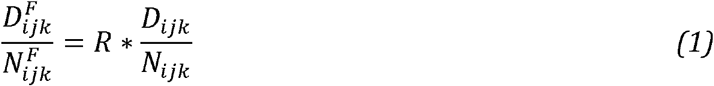

where:

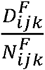 is the mortality rate among front-line workers of a given age group *i*, race/ethnicity group *j* and state *k*, and

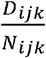 is the mortality rate among adults regardless of their occupation of that same age *i*, race/ethnicity *j* and state *k*.

Mutambudzi et al. [13] provide risk ratios for severe COVID-19 by occupational group. When compared to non-essential workers, education workers, food workers and transport workers appear to have risk ratios of respectively 1 ·56 (0 ·87-2·91), 0 ·84 (0·39-1·80) and 1·43 (0·78-2·63) in a fully adjusted model. We use 1·56 as an estimate of *R* in our main analysis, and then vary its value from 1 to 4 in sensitivity analyses, both to account for uncertainty in these estimates and variation among them, and also to account for the possibility that the value could increase during a period of restriction on economic activity, when relative exposure will be particularly high for front-line workers.

### 3.3 Estimation of the number of lives saved by a fixed number of vaccine courses *v* in a given state *k*

We aim at estimating a quantity proportional to the number of lives saved among race/ethnicity groups *j* by allocating a fixed number of vaccine courses *v* to individuals of a given age group *i* in a given US state *k*. We first describe the estimation for the purely age-based policy *S* and then describe the approach for occupation-based policy *F*. Throughout our analysis, we assume *v* = 1,000 for each state, and 95% vaccine efficacy. Because all calculations are linear in *v* and in efficacy against death, the comparative results are unaffected by the choice of values for these variables. As noted above we make the further assumption that future COVID-19 deaths in an (age, state, race) group in the absence of vaccine would be proportional to the number of deaths up to January 31, 2021. To present results, we assume that the future number of deaths in the absence of vaccine is 20% as large as the number to January 31, 2021, but again this choice has no effect on the relative magnitudes of benefit from different policies.

#### 3.3.1 Estimating the number of lives saved under policy *S*

As this policy allocated vaccines purely by age category, the number of doses allocated to each race/ethnicity group *j* is proportional to the probability that individuals in age category *i* and state *k* belong to each race/ethnicity group *j*.

Under these assumptions, the number of lives saved in a given age group *i* and race/ethnicity group *j* by *v* vaccination in state *k* is proportional to:

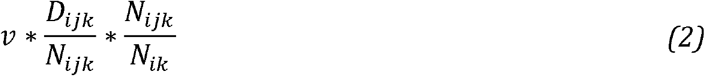

where :

*v* is the number of vaccine courses available for individuals of a given age *i* in a given state *k*,

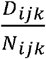 is the mortality rate among individuals of age *i*, race/ethnicity *j* in state *k*, and

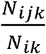 is the probability that a vaccine goes to a *j* member of the age group *i* in state *k*, assumed to be the share of *j* individuals in the age group *i*.

We can then drop the *N*_*ijk*_ term in equation 2 and write that the number of lives saved is proportional to

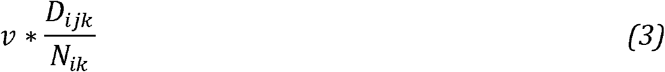

We apply this equation to estimate the number of lives saved by allocating a fixed number of vaccine courses *v* to one of two groups in a given state *k*: older individuals aged 65-74 years not in long-term care or nursing homes (policy *S*) or public-facing front-line workers aged 16-64 (policy *F*).

We’re first interested in estimating the number of lives in each race/ethnicity group *j* saved by vaccinating *v* 65-74 year old, termed *LS*_*jk*_. Applying the equations defined above in the age category 65-74 year old, we obtain:

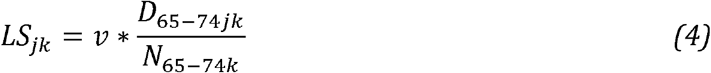

We took into account the uncertainty in the 65-74 year olds mortality rate estimate 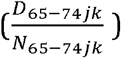 by sampling 10,000 values from a beta distribution parametrized from the number of deaths among individuals aged 65-74(*D*_65− 74*jk*_)and the number individuals aged 65-74 (*N*_65− 74*jk*_), where the α parameter is (0+ *D*_65− 74*jk*_) and the β parameter is (0+ *N*_65− 74*jk*_− *D* _65− 74*jk*_). The 10, 000 simulations of the 65-74 year olds mortality rate 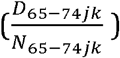 are then used to compute 10,000 values of *LS*_*jk*_, the number of lives saved under policy S. The mean, 2.5 percentile and 97.5 percentile are then computed.

#### 3.3.2 Estimating the number of lives saved under policy *F*

Next, we estimate the number of lives saved among each race/ethnicity *j* by vaccinating *v* front-line workers, termed *LF*_*jk*_.

Two layers of complexity need to be taken into account when estimating *LF*_*jk*_. First, as detailed above, deaths counts for front-line workers are not available and mortality rates among front-line workers need to be estimated from mortality rates in the general population. Second, we need to account for front-line workers having a different age distribution than the general population of adults aged 16 to 64 year old, as shown in S2 Fig. We thus start by computing *LF*_*ijk*_ among each of five age categories *i* : 16-24, 25-34, 35-44, 45-54, and 55-64 year olds.

Similarly as above,

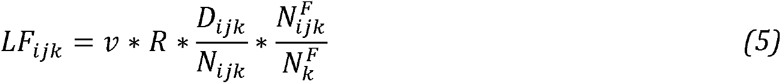

where :

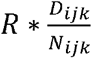 is the mortality rate among front-line workers, as detailed in section 3.2.3 above, and 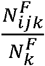 is the probability that a vaccine goes to a front-line worker of race/ethnicity *j* member of the age group *i* in state *k*, assumed to be the share of front-line workers of that age/race/ethnicity *ij* in state *k*.

We then sum all *LF*_*ijk*_ over the five age categories *i* among front-line workers to obtain the total number of lives saved for front-line workers aged 16 to 64 years, *LF*_*jk*_.

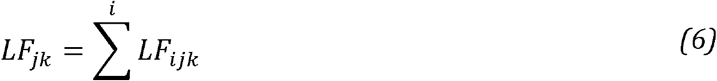

We take into account the uncertainty both in the age/race/ethnicity/state-specific mortality rate 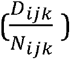, and in the proportion of front-line workers of a given age/race/ethnicity/state 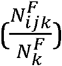 by sampling 10,000 random variables of each. For the first term, the parameters of the beta distribution are (0+ *D*_*ijk*_)and (0+ *N*_*ijk*_− *D*_*ijk*_) and for the second term the parameters of the beta distribution are 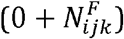 and 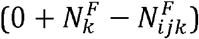. The 10,000 simulations of each of the five *LF*_*ijk*_ are summed over to obtain 10,000 values for *LF*_*jk*_, the number of lives saved under policy F. We then compute the mean, 2.5 percentile and 97.5 percentile values. We have to note that as the two terms 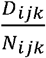 and 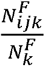 are not independent, and the resulting 95% uncertainty interval likely overestimates the dispersion in the estimated number of lives saved (*LF*_*jk*_).

#### 3.3.3 Comparing policy *S* and policy *F*

For each race/ethnicity *j* and state *k* of interest, we compute the *ratioL*_*jk*_ of the number of lives saved under each policy *S* and *F* for a given number of vaccine doses *v*. A *ratioL*_*jk*_ larger than 1 indicates that policy *S* is preferable to policy *F* for race/ethnicity *j* in state *k*.

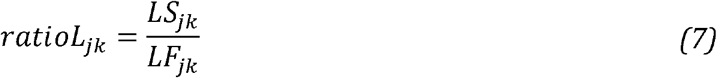

We divide each of the 10,000 estimates of *LS*_*jk*_ by one estimate of *LF*_*jk*_, thus generating 10,000 values of *ratioL*_*jk*_. These are in turn used to compute mean, 2.5 percentile and 97.5 percentile estimates.

### 3.4 Estimation of the number of years of life saved by a fixed number of vaccine courses *v* in a given state *k*

Next, we extend our analysis by estimating the number of years of life saved, instead of the number of lives.

We extend the calculations above to include an estimate of the expected number of life years saved among race/ethnicity groups. We extracted the expectations of life at different age categories between 15 and 74 years old from the WHO Global Health Observatory (GHO) [18]. As estimates are given for 5-year age bins in the WHO GHO, we average estimates two by two to obtain estimated for our 10-year age bins of interest, as shown in S2 Table. We used life expectancy estimates that do not vary by race/ethnicity groups as doing so would perpetuate existing inequities by favoring the groups that are most privileged and have the longest life expectancy, as they have most to lose.

We then adapt the calculations of the absolute number of years of life lost (YLL) described by Martinez et al. [19] to the estimation of number of years of life saved (YLS):

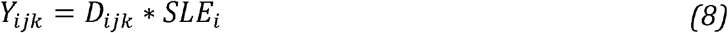

where :

*Y*_*ijk*_ is the number of years of life saved for a given age/race/ethnicity *ij* in state *k, D*_*jk*_ is the number of COVID deaths for a given age/race/ethnicity *ij* in state *k, SLE*_*i*_ is the standard life expectancy at age *i* in the USA in 2019.

We extend the equations 4, 5, and 6 above to compute *YS*_*jk*_ and *YF*_*jk*_, the number of years of lives saved under both policies *S* and *F* by including the *SLE*_*i*_ term. We thus obtain:

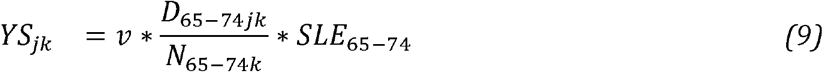

and,

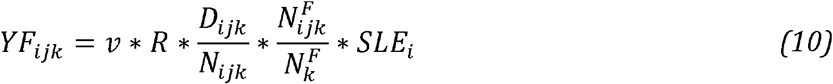

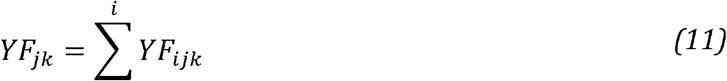

Then, similarly as in equation 7, we compute a new ratio *ratioY*_*jk*_, showing the relative performance of policies *S* and *F* in terms of number of lives saved:

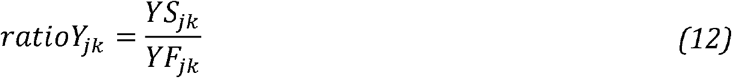

As described in section 3.3, we take into account the uncertainty by simulating a distribution of each of the input parameters. We then compute a mean and 95% interval around *YS*_*jk*_, *YF*_*jk*_ and *ratioY*_*jk*_

## 4 Results

### 4.1 Comparative vaccine dose allocation and mortality rates by race /ethnicity

Table 2 shows that Non-Hispanic Whites receive more doses under policy *S* than under policy *F*, while all other groups (Hispanics, Non-Hispanic Blacks and Non-Hispanic Asians, Non-Hispanic Two or more races, Non-Hispanic American Indian or Alaska Native, and Non-Hispanic Native Hawaiian or other Pacific Islander) receive more doses under policy *F*. This can be explained by the over-representation of Non-Hispanic Whites among elderly in the US (S1 Fig).

**Table 2.** Proportion of doses allocated under policy *S* and policy *F*, and mortality rates per 100,000 individuals for individuals 65-74 year olds and for front-line workers, per race/ethnicity categories in the United States.

| Race/ethnicity | Proportion of doses |  | Mortality rate |  |
| --- | --- | --- | --- | --- |
|  | received<br>under<br>policy S | received<br>under<br>policy F | among<br>65-74 year<br>olds | among<br>front-line<br>workers |
| All races | 100 | 100 | **39·5** | 6·9 |
| Non-Hispanic White | **74·6** | 58·2 | **27·1** | 3·7 |
| Hispanic | 9·0 | **19·7** | **95·2** | 12·6 |
| Non-Hispanic Black | 10·0 | **13·2** | **82·4** | 12·2 |
| Non-Hispanic Asian | 4·7 | **5·4** | **33·2** | 5·9 |
| Non-Hispanic 2+ races | 0·9 | **2·6** | **17·3** | 2·3 |
| Non-Hispanic Native | 0·5 | **0·7** | **105·0** | 27·5 |
| Non-Hispanic NHPI | 0·1 | **0·2** | **96·1** | 36·7 |

For each race/ethnicity category the highest proportion of doses and mortality rate are in bold. "2+ races" stands for Two or more races, "Native" stands for American Indian or Alaska Native, and "NHPI" stands for Native Hawaiian or other Pacific Islander.

Next we show the estimated COVID-19 mortality rates per 100,000 individuals, per race/ethnicity, for individuals aged 65 to 74 and for front-line workers. As shown in bold in Table 2, for all races the mortality rates are considerably higher among older individuals compared to essential workers. Also, Hispanics, Non-Hispanic Blacks, Non-Hispanic American Indian or Alaska Native and Non-Hispanic Native Hawaiian or other Pacific Islander 65-74 have higher mortality rates than Non-Hispanic Whites and Asians.

The calculations in Table 2 are repeated for each state and the District of Columbia in S3 Table.

### 4.2 Comparative number of lives saved under policies *S* and *F* by race/ethnicity

Next, we compute the number of lives saved in each race/ethnicity group by vaccinating 1,000 individuals, targeting either individuals aged 65-74 (policy *S*), or front-line workers (policy *F*). As shown in Table 3, for the total US population as well as each race/ethnicity groups, the number of lives saved is higher when vaccinating 65-74 year olds (*LS*_*j*_) compared to vaccinating front-line workers (*LF*_*j*_). The ratio 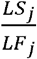 indicates for each race/ethnicity group how much better policy *S* performs when compared to policy *F*. Policy *S* is expected to save more lives for each race/ethnicity individually, and overall. However, the comparative performance of policy *S* is most beneficial for Non-Hispanic Whites than for any other race/ethnicity category.

**Table 3.**
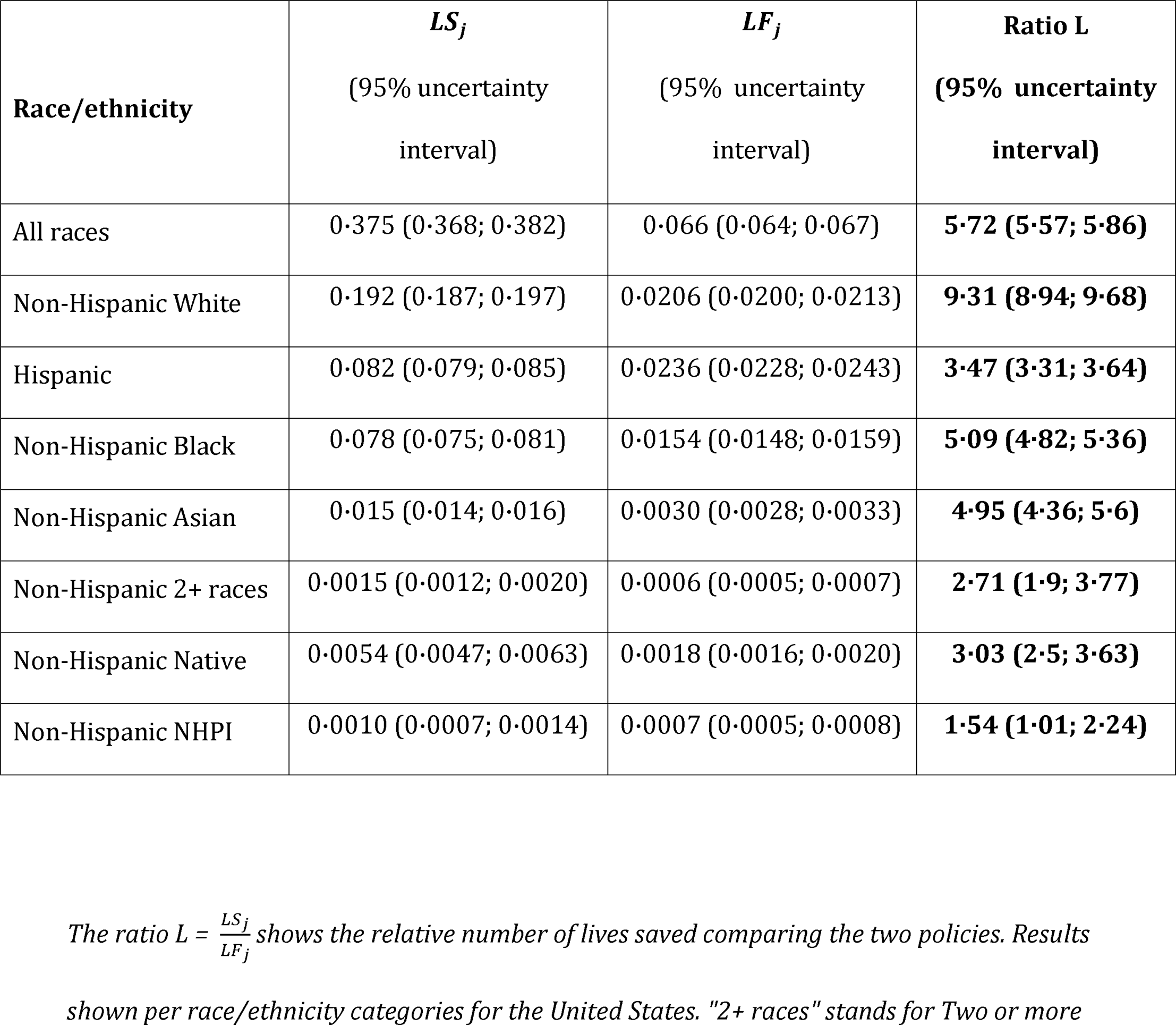
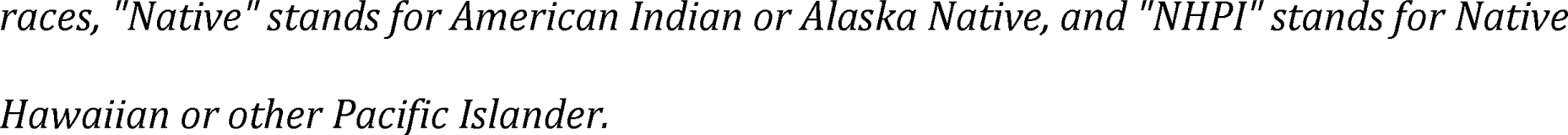
Number of lives saved in each race/ethnicity group by vaccinating 1,000 individuals, either allocating doses to individuals 65-74 (*LS*_*j*_) or to front-line workers (*LF*_*j*_).

| **Race/ethnicity** | **$LS_j$**<br>(95% uncertainty interval) | **$LF_j$**<br>(95% uncertainty interval) | **Ratio L**<br>(95% uncertainty interval) |
| --- | --- | --- | --- |
| All races | 0.375 (0.368; 0.382) | 0.066 (0.064; 0.067) | **5.72 (5.57; 5.86)** |
| Non-Hispanic White | 0.192 (0.187; 0.197) | 0.0206 (0.0200; 0.0213) | **9.31 (8.94; 9.68)** |
| Hispanic | 0.082 (0.079; 0.085) | 0.0236 (0.0228; 0.0243) | **3.47 (3.31; 3.64)** |
| Non-Hispanic Black | 0.078 (0.075; 0.081) | 0.0154 (0.0148; 0.0159) | **5.09 (4.82; 5.36)** |
| Non-Hispanic Asian | 0.015 (0.014; 0.016) | 0.0030 (0.0028; 0.0033) | **4.95 (4.36; 5.6)** |
| Non-Hispanic 2+ races | 0.0015 (0.0012; 0.0020) | 0.0006 (0.0005; 0.0007) | **2.71 (1.9; 3.77)** |
| Non-Hispanic Native | 0.0054 (0.0047; 0.0063) | 0.0018 (0.0016; 0.0020) | **3.03 (2.5; 3.63)** |
| Non-Hispanic NHPI | 0.0010 (0.0007; 0.0014) | 0.0007 (0.0005; 0.0008) | **1.54 (1.01; 2.24)** |

*The ratio $L = \frac{LS_j}{LF_j}$ shows the relative number of lives saved comparing the two policies. Results shown per race/ethnicity categories for the United States. "2+ races" stands for Two or more*
*racess, "Native" stands for American Indian or Alaska Native, and "NHPI" stands for Native Hawaiian or other Pacific Islander.*

The calculations in Table 3 are repeated for each state and the District of Columbia for which the number of deaths reported in each racial/ethnic group among both 16-64 and 65-74 year olds was larger than 2, as shown in Fig 1 and S4 Table. In all of race and state combinations for which ratios could be computed, ratios are larger than 1, suggesting that policy *S* performs best across geographies. Again, Non-Hispanic Whites benefited most from policy *S*.

**Fig 1.**
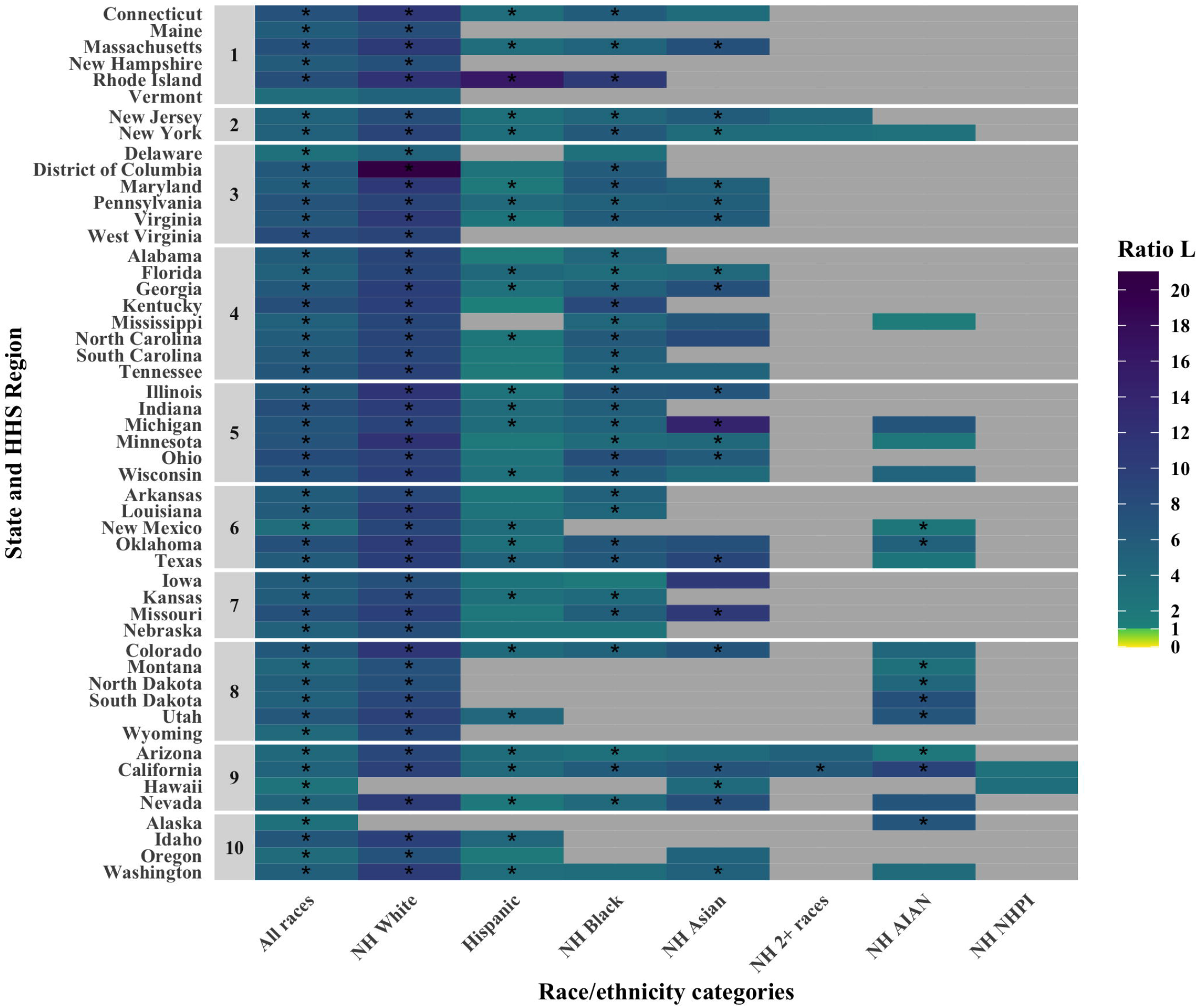
Comparative performance of policies S and F (shown in the form of a ratio 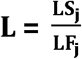) for all race/ethnicity categories in all states and the District of Columbia in terms of lives saved. States are grouped by HHS Region. Stars indicate the race/ethnicity/state for which the lower bound of the 95% interval did not include 1.

The calculation of the ratio in Table 3 are repeated for varying values of *R*, the increased risk of infection for front-line workers, as shown in S5 Table. Policy *S* performs best for all race/ethnicity categories for values of *R* up to 2·4. For *R* values between 2·5 and 4·2, policy *S* still performs best in all but one race/ethnicity category, Non-Hispanic Native Hawaiian or other Pacific Islander.

### 4.3 Comparative number of years of life saved under policies *S* and *F* by race/ethnicity

Next, we compute the number of years of life saved in each race/ethnicity group by vaccinating 1,000 individuals, targeting either individuals aged 65-74 (policy *S*), or front-line workers (policy *F*).

As shown in Table 4, for the whole US populations, as well as for most racial/ethnicity groups, more years of life are saved under policy *S* than policy *F*. It seems like only Non-Hispanic Native Hawaiian and other Pacific Islanders would benefit more from a vaccination strategy targeting front-line workers, and the uncertainty interval for this ratio overlaps with the null value 1. Similarly to Table 3, Non-Hispanic Whites would benefit most from a vaccination priorizing older 65-74 individuals.

**Table 4.** Number of years of life saved in each race/ethnicity group by vaccinating 1,000 individuals, either allocating doses to individuals 65-74 (*YS*_*j*_) or to front-line workers (*YF*_*j*_).

| Race/ethnicity | $YS_j$<br>(95% uncertainty<br>interval) | $YF_j$<br>(95% uncertainty<br>interval) | Ratio Y<br>(95% uncertainty<br>interval) |
| --- | --- | --- | --- |
| All races | 6.53 (6.42; 6.65) | 2.08 (2.04; 2.12) | **3.14 (3.06; 3.23)** |
| Non-Hispanic White | 3.35 (3.26; 3.43) | 0.62 (0.59; 0.64) | **5.44 (5.21; 5.67)** |
| Hispanic | 1.43 (1.37; 1.48) | 0.77 (0.74; 0.79) | **1.85 (1.76; 1.95)** |
| Non-Hispanic Black | 1.36 (1.31; 1.41) | 0.49 (0.47; 0.51) | **2.77 (2.62; 2.93)** |
| Non-Hispanic Asian | 0.26 (0.24; 0.28) | 0.094 (0.085; 0.102) | **2.79 (2.45; 3.16)** |
| Non-Hispanic 2+ races | 0.03 (0.02; 0.04) | 0.021 (0.017; 0.027) | **1.29 (0.88; 1.82)** |
| Non-Hispanic Native | 0.10 (0.08; 0.11) | 0.066 (0.058; 0.074) | **1.45 (1.19; 1.75)** |
| Non-Hispanic NHPI | 0.018 (0.012; 0.024) | 0.024 (0.019; 0.030) | **0.73 (0.47; 1.07)** |

*The ratio $Y = \frac{Y_{Sj}}{Y_{Fj}}$ shows the relative number of lives saved comparing the two policies. Results* *shown per race/ethnicity categories for the United States. "2+ races" stands for Two or more* *racess, "Native" stands for American Indian or Alaska Native, and "NHPI" stands for Native* *Hawaiian or other Pacific Islander.*

The ratios 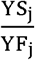 shown in Table 4 are closer to 1 than the ratios 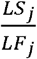 shown in Table 3, indicating that accounting for number of years of life lost leads to more similar performance for both policies.

The calculations in Table 4 are repeated for each state and the District of Columbia for which the number of deaths reported in each racial/ethnic group among both 16-64 and 65-74 year olds was larger than 2, as shown in Fig 2 and S6 Table. In the vast majority of race and state combinations for which ratios could be computed, ratios are larger than 1, suggesting that policy *S* performs best across geographies. Nine race/ethnicity/state combinations lead to ratios smaller than 1, indicative of a better performance of policy *F*, as shown in orange colours in Fig 2. However, none of these excluded 1 from the uncertainty interval.

**Fig 2.**
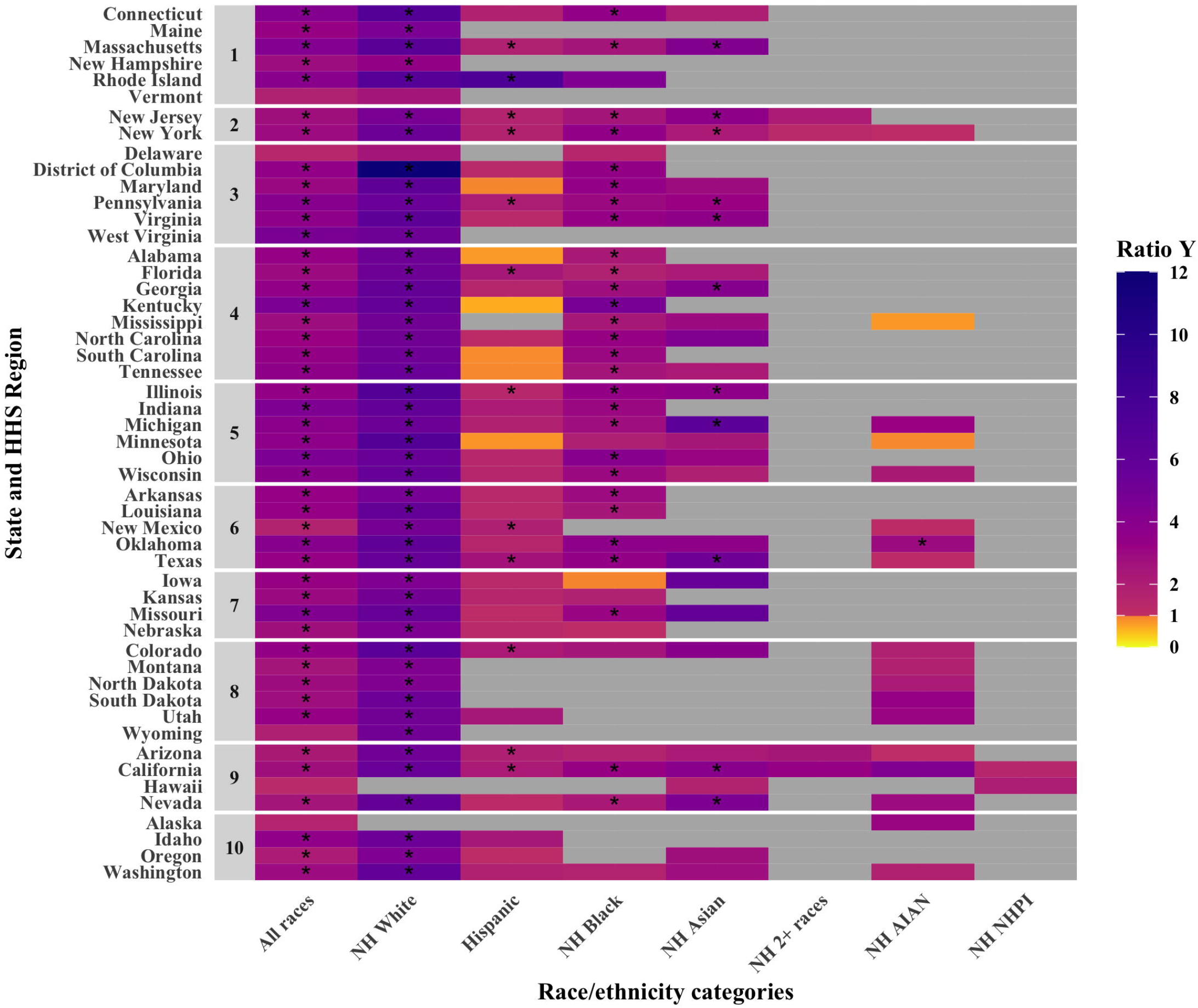
Comparative performance of policies S and F (shown in the form of a ratio Y = 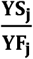) for all race/ethnicity categories in all states and the District of Columbia in terms of years of life saved. States are grouped by HHS Region. Stars indicate the race/ethnicity/state for which the lower bound of the 95% interval did not include 1.

The calculation of the ratio in Table 4 are repeated for varying values of *R*, the increased risk of infection for front-line workers, as shown in S7 Table. For values of *R* up to 2, policy *F* performs best only for Non-Hispanic Native Hawaiian or other Pacific Islander. For R values of 3 or 4, policy *F* performs better in 4 race/ethnicity groups.

## 5 Discussion

The burden of COVID-19 infections and deaths has been unequally distributed among racial and ethnic groups in the US, while deaths have been sharply higher among older age groups. The ethical imperatives of saving the most lives and minimizing inequalities have led to debate about age- and occupation-based criteria for prioritizing access to vaccines when they are scarce [20–22]. Because the hardest-hit racial and ethnic groups in the US also have younger age distributions, prioritization schemes that favor older ages intrinsically provide fewer vaccines to members of these disadvantaged groups, as we quantify in Table 2. This consideration has led some to suggest that fairness goals could be better achieved by prioritizing vaccines for front-line workers, a category that has greater exposure to infection, has high pandemic-related excess mortality [23], and that includes members of highly-affected racial and ethnic groups.

One might expect that a policy such as prioritizing front-line workers, which provides more vaccine doses to Black, Hispanic, and Asian persons than prioritizing 65-74 year olds, would also save more lives in these non-White groups. On the other hand, some have argued that this is unlikely because for each race/ethnicity, risk of death increases dramatically with age, and providing more doses to relatively young members of the hardest-hit groups ahead of (a smaller number of) older members of these groups misses the opportunity to prevent deaths in these groups. Consistent with this latter argument, we have shown that in the United States as a whole and in nearly every state, a front-line worker prioritization saves fewer lives in each racial/ethnic group than prioritization of 65-74-year-olds, at least when considering the direct effects of vaccination in protecting the vaccinated person.

Specifically, prioritizing 65-74 year olds is projected to save 3·5 times as many Hispanic lives, 5·1 times as many non-Hispanic Black, 4·9 times as many non-Hispanic Asian lives, 3·0 times as many Non-Hispanic American Indian and Alaska Native lives, 1·5 times as many Non-Hispanic Native Hawaiian or other Pacific Islander, 2·7 times as many non-Hispanic multiracial lives, and 9·4 times more non-Hispanic White lives than prioritizing front-line workers. Simply put, this is because the dramatically higher COVID-19 mortality rates in older persons (ranging from 2·6 to 7·5 fold by our estimates for the different race/ethnicity groups) outweigh the more modest differences in representation of these groups among different races (ranging from 1·1-fold for Asians to 2·8-fold for individuals with two or more races). This is likely driven by the over-representation of younger adults among the front-line workers when compared to the general population (S2 Fig). In short, Blacks, Hispanics, Asians, American Indians and Alaska Natives, Native Hawaiian and other Pacific Islanders and individuals with more than one race get somewhat more vaccines under a front-line worker priority, but those vaccines go to individuals at much lower risk of dying.

This finding demonstrates by example that it is possible to choose a vaccine distribution strategy that provides a more equal vaccine distribution by race/ethnicity, yet prevents fewer deaths in every racial/ethnic group. Under our assumptions, if one considers vaccines as means to the ends of saving lives, the greater number of lives saved under the age-based policy, including in the hardest-hit groups, should lead one to favor the age-based policy over the front-line-worker policy regardless of how one weighs overall societal benefit and benefit to the hardest-hit groups, because the age-based policy maximizes both.

Our model relies on multiple simplifying modelling assumptions. We assume that (i) the vaccine efficacy and uptake is constant across age, race and location, (ii) the proportion of deaths that took place in nursing homes and long-term care facilities is constant across races for a given age and state (an assumption made because nursing home status of deaths is given by age and state, but not race), (iii) 10% of deaths recorded for the 15-24 year old category took place among 15 year olds, (iv) that we can account for the increased risk of infection and deaths among front-line workers by multiplying the mortality rate by an increased risk *R* constant across age, race and state, and that (v) analyses by Mutambudzi et al. [13] appropriately estimate this *R*.

We have shown in sensitivity analyses that the increase in infection risk for front-line workers would need to be larger than 3 for a policy vaccinating front-line workers over 65-74 year olds to become preferable for some race/ethnicity groups (S5 and S7 Table). Such large values would be more likely to happen during lock-down periods, when public-facing workers have much larger contact rates than the rest of the population. If vaccines are being rolled-out during such periods, then a policy allocating doses to front-line workers first may be preferable.

Our analysis is subject to other limitations. First, we focus our analysis on two outcomes: deaths and years of life lost. We did not assess the impact of the two vaccination policies on other outcomes such as number of infections, morbidity such as ‘long COVID’, hospitalization or other economic consequences. As it has been show that vaccination strategies targeting younger adults can lead to lower number of infections but higher number of deaths [9], it is possible that vaccinating front-line workers would lead to less infections. Second, we assume that the number of deaths occurring after vaccine allocation and roll-out is proportional to the death count up to January 30, 2021 (more exactly is it assumed to be equal to 20% of the cumulative deaths up to that date in each age, race and state combination). We thus assume that the inequalities in infection and mortality rates stay constant over the duration of the pandemic. Third, we only consider the direct effect of vaccination on the vaccinated, and not the indirect effect of vaccination protecting the rest of the population by limiting onward transmission. When this work began, it was unclear how to what extent COVID-19 vaccines reduce transmission [24]. While evidence is now accumulating for substantial vaccine effects on infection and onward spread [25], these results reflect the knowledge available at the time prioritization decisions were being made. Moreover, as the B.1.617.2 (Delta) and B.1.1.529 (Omicron) variants have become dominant worldwide, doubts on vaccine efficacy against it are being raised [26]. If the protection against transmission of Delta, Omicron, and future variants is indeed found to be limited, vaccine allocation strategies should focus on direct protection from infection. The magnitude of the relative benefit of the 65-74 strategy over the front-line strategy via direct protection studied here suggests that the greater indirect effects of vaccinating front-line workers would have to be large to make it superior in preventing deaths in any racial/ethnic group, or overall. Front-line workers and members of racial/ethnic minorities [27] may be more likely to live in multi-generation households and contribute to more onward transmission than 65-74 year old individuals. Mulberry et al. [28] studied the indirect effect of vaccination (without considering race/ ethnicity) using an age-stratified SEIR model including compartment for essential workers. Under strong assumptions that drastic social distancing measured are in place (no workplace contacts for non-essential workers and all contacts reduced), that the impact of vaccination of transmission is strong, and that *R*_*t*_ is not kept at very low levels, they showed that vaccinating essential workers and/or all adults 20-79 after individuals over 80 leads to lower number of infections, hospitalization and deaths than a solely age-based strategy. Further work is needed to understand these tradeoffs and the extent to which accounting for indirect protection might reverse our findings under the rapidly changing conditions of COVID-19.

Notwithstanding these limitations, we have shown that from the perspective of infections directly prevented, a front-line-worker COVID-19 vaccine prioritization strategy does not save as many lives and years of life as prioritization of 65-74 year olds, overall or in each racial/ethnic group, and that this effect is qualitatively consistent across US states.

Although these two strategies received the most attention from committees considering prioritization schemes, there may be other strategies that might outperform either one, as some work has already suggested. For example, once health-care workers and individuals aged 65+ are vaccinated, prioritizing adults at increased risk of severe COVID-19 would increase access to vaccines for Non-Hispanic Blacks, while Hispanics would receive most doses under a strategy targeting essential workers and Non-Hispanic Whites and Asians under a solely age-based strategy [29]. For the goal of saving the most lives with a prevention intervention such as vaccination, it is optimal to prioritize individuals in order of their estimated mortality risk from the disease, which is the product of the mortality risk if infected (the infection-fatality risk) and their probability of becoming infected (a function of exposure). It is evident in the United States and elsewhere, at least from the pandemic history to date, that both older age and non-White race or Hispanic ethnicity are associated with higher risks [30]. Mortality risk at a given age varies with racial/ethnic classification, as for instance, in the US in 2020 Non-Hispanic American Indian or Alaskan Native aged 35-44 and Hispanic individuals aged 45-54 had higher mortality rates than Non-Hispanic White individuals aged 55-64 (S3 Fig). It follows that a policy that allocated vaccine priority according to this criterion would at each age prioritize Black and Hispanic individuals over White ones, and would assign equal priority, all other factors being equal, to a younger Hispanic or Black individual as to an older White one [31]. A version of this approach – lower age cutoffs for Black persons to receive priority – has been proposed [32] and would both enhance equity and, if done appropriately, also enhance the number of lives saved overall and for Black Americans compared to an age-only prioritization. Likewise, geographic targeting to areas of high risk would increase racial equity while improving access to vaccination for those at greater risk of death from COVID-19 [31]. Overall, questions related to the fairness of vaccine prioritization are most important when vaccine distribution is slow. If the roll-out is rapid, the difference in the benefit from different strategies diminishes, as all individuals get access to vaccines at similar times regardless of their age, race/ethnicity and occupation.

## Data Availability

This study involves only openly available data obtained from the American Community Survey (ACS) and the National Center for Health Statistics (NCHS), which can be obtained from https://data.cdc.gov/NCHS/Deaths-involving-coronavirus-disease-2019-COVID-19/ks3g-spdg, https://data.cdc.gov/NCHS/NVSS-Provisional-COVID-19-Deaths-by-Place-of-Death/4va6-ph5s, and https://www.census.gov/programs-surveys/acs/data.html.

https://data.cdc.gov/NCHS/NVSS-Provisional-COVID-19-Deaths-by-Place-of-Death/4va6-ph5s

https://www.census.gov/programs-surveys/acs/data.html

## 6 Acknowledgement

The authors thank Dr. Elizabeth Wrigley-Field and Dr. Ayesha Mahmud for useful comments on an earlier version of this work.

## 8 Supplementary Information

**S1 Fig. Proportion of adults aged 16-64, front-line workers aged 16-64, and older individuals aged 65-74 in each race/ethnicity categories**.

**S2 Fig. Proportion of adults and front-line workers in each age subcategories between 16 and 64**.

**S3 Fig. COVID-19 mortality rates per 100**,**000 by age and race/ethnicity categories in the US in 2020**.

**S1 Table. List of occupations included in the definition of front-line occupations. For each race, proportion of front-line workers in each occupation**.

**S2 Table. Expectation of life at age categories i9/27/2022 4:14:00 PMin the USA in 2019 from the WHO GlobalHealth Observatory [18]**.

**S3 Table. Proportion of doses allocated under policy S and policy F, and mortality rates per100**,**000 individuals for individuals 65-74 year olds and for front-line workers, per US state and race/ethnicity categories**. “NH” stands for “Non-Hispanic”, “2+ races” stands for “Two or more races”, “Native” stands for “American Indian or Alaska Native”, “NHPI” stands for “Native Hawaiian or Other Pacific Islander”.

**S4 Table. Number of lives saved by vaccinating 1**,**000 individuals, either allocating doses to 65-74 year olds (policy S), or to front line workers (policy F). The ratio** 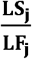 **shows the relative number of lives saved comparing the two policies. Results shown for each US state per race/ethnicity categories**. “NH” stands for “Non-Hispanic”, “2+ races” stands for “Two or more races”, “Native” stands for “American Indian or Alaska Native”, “NHPI” stands for “Native Hawaiian or Other Pacific Islander”.

**S5 Table. Relative number of lives saved comparing a policy allocating vaccines to 65-74 year olds (policy S) to one allocating vaccines to front-line workers (policy F). Results shown for varying values of *R*, the increased risk of infection for front-line workers**. The ratios for an R value of 1·56 correspond to those shown in Table 3. “NH” stands for “Non-Hispanic”, “2+ races” stands for “Two or more races”, “Native” stands for “American Indian or Alaska Native”, “NHPI” stands for “Native Hawaiian or Other Pacific Islander”.

**S6 Table. Number of years of life saved by vaccinating 1**,**000 individuals, either allocating doses to 65-74 year olds (policy S), or to front line workers (policy F). The ratio** 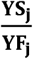 **shows the relative number of lives saved comparing the two policies. Results shown for each US state per race/ethnicity categories**. “NH” stands for “Non-Hispanic”, “2+ races” stands for “Two or more races”, “Native” stands for “American Indian or Alaska Native”, “NHPI” stands for “Native Hawaiian or Other Pacific Islander”.

**S7 Table. Relative number of years of life saved comparing a policy allocating vaccines to 65-74 year olds (policy S) to one allocating vaccines to front-line workers (policy F). Results shown for varying values of *R*, the increased risk of infection for front-line workers**. The ratios for an R value of 1·56 correspond to those shown in Table 4. “NH” stands for “Non-Hispanic”, “2+ races” stands for “Two or more races”, “Native” stands for “American Indian or Alaska Native”, “NHPI” stands for “Native Hawaiian or Other Pacific Islander”.

