## Supplementary material for "Fairness and efficiency considerations in COVID-19 vaccine allocation strategies: a case study comparing front-line workers and 65-74 year olds in the United States": 2022-09-12-Vaccine_prioritization_Supplementary_Table1.docx

**S1 Table. List of occupations included in the definition of front-line occupations. For each race, proportion of front-line workers in each occupation.**

| Occupation | Category | All races | Non-Hispanic White | Hispanic | Non-Hispanic Black | Non-Hispanic Asian | Non-Hispanic Two or more races | Non-Hispanic American Indian or Alaska Native | Non-Hispanic Native Hawaiian or other Pacific Islander |
| --- | --- | --- | --- | --- | --- | --- | --- | --- | --- |
| Cashiers | Sales | 14.01 | 11.72 | 16.30 | 19.11 | 15.76 | 16.27 | 18.95 | 21.36 |
| Retail Salespersons | Sales | 12.25 | 12.41 | 12.20 | 11.57 | 12.00 | 13.99 | 9.37 | 8.50 |
| First-Line Supervisors Of Retail Sales Workers | Sales | 10.47 | 11.89 | 8.18 | 7.73 | 11.42 | 8.48 | 9.11 | 9.07 |
| Elementary And Middle School Teachers | Education | 10.43 | 13.21 | 5.68 | 7.98 | 6.27 | 6.15 | 8.07 | 3.82 |
| Cooks | Food industry | 9.56 | 7.15 | 14.62 | 12.77 | 8.86 | 9.13 | 14.07 | 10.28 |
| Waiters And Waitresses | Food industry | 8.93 | 8.99 | 9.73 | 6.19 | 11.41 | 10.50 | 8.74 | 8.67 |
| Secondary School Teachers | Education | 4.31 | 5.83 | 2.01 | 2.12 | 2.56 | 3.25 | 2.69 | 1.29 |
| Food Preparation Workers | Food industry | 4.22 | 3.65 | 5.62 | 4.24 | 4.65 | 4.78 | 5.60 | 5.75 |
| Preschool And Kindergarten Teachers | Education | 3.17 | 3.54 | 2.34 | 3.19 | 2.37 | 2.77 | 2.82 | 3.53 |
| Fast Food And Counter Workers | Food industry | 3.03 | 2.94 | 3.02 | 3.12 | 3.20 | 4.48 | 2.55 | 4.47 |
| First-Line Supervisors Of Food Preparation And Serving Workers | Food industry | 2.02 | 1.98 | 2.14 | 2.14 | 1.48 | 2.42 | 1.70 | 1.73 |
| Chefs And Head Cooks | Food industry | 1.86 | 1.40 | 2.16 | 1.83 | 5.85 | 1.69 | 1.22 | 3.79 |
| Bartenders | Food industry | 1.74 | 2.20 | 1.32 | 0.66 | 0.86 | 1.94 | 1.67 | 0.46 |
| Taxi Drivers | Transportation | 1.71 | 1.03 | 2.14 | 3.14 | 4.29 | 1.66 | 0.73 | 0.92 |
| Dishwashers | Food industry | 1.60 | 1.26 | 2.15 | 2.29 | 1.01 | 2.09 | 3.79 | 2.72 |
| Hosts And Hostesses, Restaurant, Lounge, And Coffee Shop | Food industry | 1.49 | 1.53 | 1.55 | 1.14 | 1.22 | 2.57 | 0.96 | 1.34 |
| Special Education Teachers | Education | 1.47 | 1.94 | 0.70 | 1.09 | 0.53 | 0.87 | 1.17 | 0.50 |
| Dining Room And Cafeteria Attendants And Bartender Helpers | Food industry | 1.40 | 1.20 | 2.16 | 1.21 | 1.27 | 1.35 | 1.22 | 0.74 |
| Bus Drivers, School | Transportation | 0.91 | 0.94 | 0.67 | 1.48 | 0.25 | 0.46 | 1.11 | 1.40 |
| Butchers And Other Meat, Poultry, And Fish Processing Workers | Food industry | 0.85 | 0.63 | 1.39 | 1.19 | 0.62 | 0.41 | 0.66 | 2.30 |
| Bus Drivers, Transit And Intercity | Transportation | 0.83 | 0.51 | 0.83 | 2.36 | 0.57 | 0.71 | 0.79 | 1.59 |
| Food Servers, Nonrestaurant | Food industry | 0.74 | 0.65 | 0.64 | 1.10 | 1.13 | 0.88 | 0.53 | 2.14 |
| Aircraft Pilots And Flight Engineers | Transportation | 0.68 | 1.00 | 0.23 | 0.13 | 0.29 | 0.75 | 0.23 | 0.98 |
| Door-To-Door Sales Workers, News And Street Vendors, And Related Workers | Sales | 0.46 | 0.48 | 0.53 | 0.32 | 0.26 | 0.57 | 0.43 | 0.29 |
| Parts Salespersons | Sales | 0.40 | 0.50 | 0.37 | 0.18 | 0.11 | 0.27 | 0.15 | 0.29 |
| Flight Attendants | Transportation | 0.38 | 0.39 | 0.26 | 0.41 | 0.58 | 0.59 | 0.15 | 0.44 |
| Counter And Rental Clerks | Sales | 0.34 | 0.34 | 0.37 | 0.31 | 0.35 | 0.18 | 0.69 | 0.80 |
| Shuttle Drivers And Chauffeurs | Transportation | 0.33 | 0.22 | 0.40 | 0.60 | 0.62 | 0.32 | 0.31 | 0.26 |
| Air Traffic Controllers And Airfield Operations Specialists | Transportation | 0.15 | 0.18 | 0.11 | 0.10 | 0.05 | 0.27 | 0.02 | NA |
| Morticians, Undertakers, And Funeral Arrangers | Funeral workers | 0.12 | 0.15 | 0.05 | 0.13 | 0.06 | 0.09 | 0.01 | 0.22 |
| Food Preparation and Serving Related Workers, All Other | Food industry | 0.06 | 0.04 | 0.08 | 0.06 | 0.12 | 0.07 | 0.17 | 0.06 |
| Embalmers, Crematory Operators And Funeral Attendants | Funeral workers | 0.04 | 0.05 | 0.03 | 0.05 | 0.01 | 0.01 | 0.03 | 0.27 |
| Ambulance Drivers And Attendants, Except Emergency Medical Technicians | Transportation | 0.03 | 0.03 | 0.01 | 0.03 | 0.01 | 0.01 | 0.30 | NA |
