## Supplementary material for "Fairness and efficiency considerations in COVID-19 vaccine allocation strategies: a case study comparing front-line workers and 65-74 year olds in the United States": 2022-09-12-Vaccine_prioritization_Supplementary_Table2.docx

**S2 Table. Expectation of life at age categories i in the USA in 2019 from the WHO GlobalHealth Observatory [18].**

| **Age category** | **SLE (years)** |
| --- | --- |
| 15-24 | 61.67 |
| 25-34 | 52.15 |
| 35-44 | 42.84 |
| 45-54 | 33.70 |
| 55-64 | 25.14 |
| 65-74 | 17.42 |
