## Supplementary material for "Fairness and efficiency considerations in COVID-19 vaccine allocation strategies: a case study comparing front-line workers and 65-74 year olds in the United States": 2022-09-12-Vaccine_prioritization_Supplementary_Table3.docx

**S3 Table. Proportion of doses allocated under policy S and policy F, and mortality rates per 100,000 individuals for individuals 65-74 year olds and for front-line workers, per US state and race/ethnicity categories.**

"NH" stands for "Non-Hispanic", "2+ races" stands for "Two or more races", "Native" stands for "American Indian or Alaska Native", "NHPI" stands for "Native Hawaiian or Other Pacific Islander".

| **State** | **Race/ ethnicity** | **Proportion of doses received under policy S** | **Proportion of doses received under policy F** | **Mortality rate among 65-74 year olds** | **Mortality rate among front-line workers** |
| --- | --- | --- | --- | --- | --- |
| Alabama | All races | 1.00 | 1.00 | 53.19 | 9.01 |
| Alabama | NH White | 0.74 | 0.61 | 44.76 | 6.09 |
| Alabama | Hispanic | 0.01 | 0.05 | 58.04 | 12.07 |
| Alabama | NH Black | 0.22 | 0.31 | 86.25 | 14.47 |
| Alabama | NH Asian | 0.01 | 0.01 | 17.21 | 12.63 |
| Alabama | NH 2+ races | 0.01 | 0.02 | 0.00 | 3.32 |
| Alabama | NH Native | 0.00 | 0.00 | 44.03 | 37.22 |
| Alabama | NH NHPI | 0.00 | 0.00 | 0.00 | 0.00 |
| Alaska | All races | 1.00 | 1.00 | 13.63 | 4.58 |
| Alaska | NH White | 0.73 | 0.64 | 4.17 | 0.77 |
| Alaska | Hispanic | 0.04 | 0.07 | 0.00 | 2.03 |
| Alaska | NH Black | 0.01 | 0.04 | 96.06 | 5.90 |
| Alaska | NH Asian | 0.07 | 0.06 | 19.66 | 14.68 |
| Alaska | NH 2+ races | 0.02 | 0.10 | 72.74 | 2.95 |
| Alaska | NH Native | 0.09 | 0.09 | 54.24 | 10.16 |
| Alaska | NH NHPI | 0.03 | 0.01 | 51.39 | 142.92 |
| Arizona | All races | 1.00 | 1.00 | 45.31 | 10.86 |
| Arizona | NH White | 0.77 | 0.54 | 25.91 | 4.30 |
| Arizona | Hispanic | 0.14 | 0.31 | 117.78 | 15.09 |
| Arizona | NH Black | 0.03 | 0.05 | 60.59 | 10.74 |
| Arizona | NH Asian | 0.02 | 0.03 | 33.52 | 6.67 |
| Arizona | NH 2+ races | 0.01 | 0.02 | 57.01 | 4.67 |
| Arizona | NH Native | 0.02 | 0.04 | 217.62 | 64.83 |
| Arizona | NH NHPI | 0.00 | 0.00 | 148.07 | 45.21 |
| Arkansas | All races | 1.00 | 1.00 | 41.42 | 7.06 |
| Arkansas | NH White | 0.83 | 0.72 | 35.84 | 5.03 |
| Arkansas | Hispanic | 0.02 | 0.07 | 112.61 | 11.76 |
| Arkansas | NH Black | 0.13 | 0.15 | 59.06 | 9.71 |
| Arkansas | NH Asian | 0.01 | 0.02 | 108.54 | 14.29 |
| Arkansas | NH 2+ races | 0.01 | 0.02 | 26.70 | 7.03 |
| Arkansas | NH Native | 0.00 | 0.00 | 62.26 | 11.89 |
| Arkansas | NH NHPI | 0.00 | 0.01 | NA | NA |
| California | All races | 1.00 | 1.00 | 29.70 | 5.86 |
| California | NH White | 0.55 | 0.35 | 14.09 | 2.32 |
| California | Hispanic | 0.22 | 0.42 | 73.79 | 9.55 |
| California | NH Black | 0.05 | 0.05 | 42.72 | 7.29 |
| California | NH Asian | 0.16 | 0.13 | 18.25 | 3.24 |
| California | NH 2+ races | 0.01 | 0.03 | 15.03 | 1.29 |
| California | NH Native | 0.00 | 0.00 | 45.70 | 7.25 |
| California | NH NHPI | 0.00 | 0.00 | 74.98 | 21.21 |
| Colorado | All races | 1.00 | 1.00 | 24.82 | 3.95 |
| Colorado | NH White | 0.82 | 0.66 | 17.17 | 1.98 |
| Colorado | Hispanic | 0.11 | 0.22 | 66.35 | 8.76 |
| Colorado | NH Black | 0.03 | 0.04 | 59.12 | 7.95 |
| Colorado | NH Asian | 0.02 | 0.04 | 44.15 | 4.83 |
| Colorado | NH 2+ races | 0.01 | 0.03 | 14.54 | 2.29 |
| Colorado | NH Native | 0.00 | 0.01 | 95.11 | 22.97 |
| Colorado | NH NHPI | 0.00 | 0.00 | 193.81 | 24.83 |
| Connecticut | All races | 1.00 | 1.00 | 38.73 | 5.41 |
| Connecticut | NH White | 0.82 | 0.63 | 30.19 | 3.61 |
| Connecticut | Hispanic | 0.08 | 0.20 | 71.40 | 8.23 |
| Connecticut | NH Black | 0.07 | 0.11 | 114.46 | 12.96 |
| Connecticut | NH Asian | 0.03 | 0.05 | 19.94 | 4.43 |
| Connecticut | NH 2+ races | 0.01 | 0.02 | 0.00 | 0.00 |
| Connecticut | NH Native | 0.00 | 0.00 | 67.85 | 0.00 |
| Connecticut | NH NHPI | 0.00 | 0.00 | 0.00 | NA |
| Delaware | All races | 1.00 | 1.00 | 21.57 | 7.35 |
| Delaware | NH White | 0.78 | 0.62 | 17.92 | 4.88 |
| Delaware | Hispanic | 0.03 | 0.10 | 49.51 | 13.22 |
| Delaware | NH Black | 0.15 | 0.20 | 39.58 | 10.38 |
| Delaware | NH Asian | 0.03 | 0.05 | 0.00 | 7.10 |
| Delaware | NH 2+ races | 0.00 | 0.03 | 0.00 | 3.61 |
| Delaware | NH Native | 0.00 | 0.00 | 0.00 | 169.49 |
| Delaware | NH NHPI | 0.00 | 0.00 | NA | NA |
| District of Columbia | All races | 1.00 | 1.00 | 94.56 | 15.11 |
| District of Columbia | NH White | 0.34 | 0.22 | 22.64 | 2.26 |
| District of Columbia | Hispanic | 0.07 | 0.17 | 185.40 | 30.03 |
| District of Columbia | NH Black | 0.54 | 0.56 | 128.51 | 21.40 |
| District of Columbia | NH Asian | 0.03 | 0.03 | 58.45 | 9.08 |
| District of Columbia | NH 2+ races | 0.01 | 0.02 | 176.92 | 0.00 |
| District of Columbia | NH Native | 0.00 | 0.00 | 0.00 | 343.27 |
| District of Columbia | NH NHPI | 0.00 | 0.00 | NA | NA |
| Florida | All races | 1.00 | 1.00 | 35.18 | 6.68 |
| Florida | NH White | 0.71 | 0.50 | 23.74 | 3.91 |
| Florida | Hispanic | 0.16 | 0.27 | 59.59 | 8.31 |
| Florida | NH Black | 0.10 | 0.18 | 82.82 | 13.42 |
| Florida | NH Asian | 0.02 | 0.03 | 25.28 | 5.56 |
| Florida | NH 2+ races | 0.01 | 0.02 | 12.24 | 2.39 |
| Florida | NH Native | 0.00 | 0.00 | 19.82 | 21.12 |
| Florida | NH NHPI | 0.00 | 0.00 | 95.51 | 34.93 |
| Georgia | All races | 1.00 | 1.00 | 44.72 | 7.10 |
| Georgia | NH White | 0.67 | 0.50 | 33.28 | 4.59 |
| Georgia | Hispanic | 0.03 | 0.09 | 83.79 | 10.74 |
| Georgia | NH Black | 0.26 | 0.36 | 71.84 | 10.10 |
| Georgia | NH Asian | 0.03 | 0.04 | 29.88 | 3.63 |
| Georgia | NH 2+ races | 0.01 | 0.02 | 12.85 | 2.64 |
| Georgia | NH Native | 0.00 | 0.00 | 0.00 | 14.53 |
| Georgia | NH NHPI | 0.00 | 0.00 | 144.85 | 0.00 |
| Hawaii | All races | 1.00 | 1.00 | 9.86 | 3.65 |
| Hawaii | NH White | 0.30 | 0.19 | 2.04 | 1.37 |
| Hawaii | Hispanic | 0.03 | 0.11 | 18.78 | 6.23 |
| Hawaii | NH Black | 0.01 | 0.01 | 67.32 | 13.66 |
| Hawaii | NH Asian | 0.47 | 0.39 | 8.96 | 3.13 |
| Hawaii | NH 2+ races | 0.11 | 0.18 | 5.73 | 2.15 |
| Hawaii | NH Native | 0.00 | 0.00 | 0.00 | 0.00 |
| Hawaii | NH NHPI | 0.08 | 0.11 | 40.71 | 10.30 |
| Idaho | All races | 1.00 | 1.00 | 27.82 | 4.33 |
| Idaho | NH White | 0.94 | 0.81 | 23.15 | 2.97 |
| Idaho | Hispanic | 0.04 | 0.14 | 116.98 | 7.94 |
| Idaho | NH Black | 0.00 | 0.01 | 2304.19 | 21.49 |
| Idaho | NH Asian | 0.01 | 0.01 | 66.57 | 23.22 |
| Idaho | NH 2+ races | 0.01 | 0.02 | 36.07 | 7.73 |
| Idaho | NH Native | 0.01 | 0.01 | 69.01 | 44.22 |
| Idaho | NH NHPI | 0.00 | 0.00 | NA | 0.00 |
| Illinois | All races | 1.00 | 1.00 | 45.48 | 7.46 |
| Illinois | NH White | 0.74 | 0.58 | 31.36 | 3.48 |
| Illinois | Hispanic | 0.08 | 0.21 | 113.39 | 15.68 |
| Illinois | NH Black | 0.13 | 0.13 | 88.49 | 12.88 |
| Illinois | NH Asian | 0.05 | 0.05 | 38.76 | 5.89 |
| Illinois | NH 2+ races | 0.01 | 0.02 | 14.81 | 3.81 |
| Illinois | NH Native | 0.00 | 0.00 | 104.39 | 23.43 |
| Illinois | NH NHPI | 0.00 | 0.00 | NA | 0.00 |
| Indiana | All races | 1.00 | 1.00 | 40.50 | 5.21 |
| Indiana | NH White | 0.89 | 0.78 | 36.02 | 4.14 |
| Indiana | Hispanic | 0.02 | 0.08 | 93.59 | 7.54 |
| Indiana | NH Black | 0.07 | 0.09 | 83.19 | 11.74 |
| Indiana | NH Asian | 0.01 | 0.02 | 26.18 | 3.83 |
| Indiana | NH 2+ races | 0.01 | 0.02 | 18.59 | 2.49 |
| Indiana | NH Native | 0.00 | 0.00 | 70.01 | 21.07 |
| Indiana | NH NHPI | 0.00 | 0.00 | 219.54 | NA |
| Iowa | All races | 1.00 | 1.00 | 35.95 | 6.29 |
| Iowa | NH White | 0.95 | 0.84 | 33.18 | 5.06 |
| Iowa | Hispanic | 0.01 | 0.06 | 99.08 | 9.69 |
| Iowa | NH Black | 0.02 | 0.05 | 84.46 | 16.58 |
| Iowa | NH Asian | 0.01 | 0.03 | 80.21 | 4.49 |
| Iowa | NH 2+ races | 0.01 | 0.01 | 42.86 | 7.13 |
| Iowa | NH Native | 0.00 | 0.00 | 198.90 | 106.23 |
| Iowa | NH NHPI | 0.00 | 0.00 | 1294.54 | NA |
| Kansas | All races | 1.00 | 1.00 | 32.44 | 5.70 |
| Kansas | NH White | 0.87 | 0.72 | 27.59 | 3.93 |
| Kansas | Hispanic | 0.05 | 0.13 | 84.59 | 10.28 |
| Kansas | NH Black | 0.05 | 0.06 | 50.44 | 12.21 |
| Kansas | NH Asian | 0.02 | 0.04 | 39.91 | 6.43 |
| Kansas | NH 2+ races | 0.01 | 0.04 | 34.42 | 2.77 |
| Kansas | NH Native | 0.01 | 0.01 | 107.70 | 43.20 |
| Kansas | NH NHPI | 0.00 | 0.00 | NA | NA |
| Kentucky | All races | 1.00 | 1.00 | 30.25 | 3.87 |
| Kentucky | NH White | 0.89 | 0.83 | 28.01 | 3.24 |
| Kentucky | Hispanic | 0.01 | 0.04 | 48.67 | 12.73 |
| Kentucky | NH Black | 0.07 | 0.09 | 57.12 | 6.12 |
| Kentucky | NH Asian | 0.01 | 0.02 | 21.63 | 10.31 |
| Kentucky | NH 2+ races | 0.01 | 0.02 | 0.00 | 0.00 |
| Kentucky | NH Native | 0.00 | 0.00 | 141.86 | 0.00 |
| Kentucky | NH NHPI | 0.00 | 0.00 | 0.00 | 0.00 |
| Louisiana | All races | 1.00 | 1.00 | 62.65 | 10.46 |
| Louisiana | NH White | 0.68 | 0.53 | 44.08 | 5.58 |
| Louisiana | Hispanic | 0.03 | 0.06 | 65.91 | 12.53 |
| Louisiana | NH Black | 0.27 | 0.37 | 111.50 | 18.58 |
| Louisiana | NH Asian | 0.01 | 0.02 | 16.70 | 11.58 |
| Louisiana | NH 2+ races | 0.01 | 0.02 | 25.93 | 0.00 |
| Louisiana | NH Native | 0.00 | 0.00 | 51.55 | 30.88 |
| Louisiana | NH NHPI | 0.00 | 0.00 | NA | NA |
| Maine | All races | 1.00 | 1.00 | 6.38 | 1.23 |
| Maine | NH White | 0.97 | 0.91 | 5.36 | 0.87 |
| Maine | Hispanic | 0.01 | 0.03 | 62.16 | 0.00 |
| Maine | NH Black | 0.01 | 0.01 | 77.83 | 32.56 |
| Maine | NH Asian | 0.01 | 0.03 | 0.00 | 0.00 |
| Maine | NH 2+ races | 0.00 | 0.02 | 87.37 | 0.00 |
| Maine | NH Native | 0.01 | 0.01 | 0.00 | 0.00 |
| Maine | NH NHPI | 0.00 | 0.00 | NA | NA |
| Maryland | All races | 1.00 | 1.00 | 37.37 | 6.58 |
| Maryland | NH White | 0.62 | 0.48 | 23.28 | 2.81 |
| Maryland | Hispanic | 0.04 | 0.11 | 85.40 | 18.49 |
| Maryland | NH Black | 0.26 | 0.31 | 66.48 | 9.16 |
| Maryland | NH Asian | 0.06 | 0.07 | 26.28 | 4.88 |
| Maryland | NH 2+ races | 0.01 | 0.03 | 11.87 | 1.82 |
| Maryland | NH Native | 0.00 | 0.00 | 49.31 | 0.00 |
| Maryland | NH NHPI | 0.00 | 0.00 | NA | NA |
| Massachusetts | All races | 1.00 | 1.00 | 34.49 | 4.68 |
| Massachusetts | NH White | 0.85 | 0.68 | 29.96 | 3.59 |
| Massachusetts | Hispanic | 0.06 | 0.15 | 65.38 | 7.01 |
| Massachusetts | NH Black | 0.05 | 0.08 | 81.85 | 10.68 |
| Massachusetts | NH Asian | 0.04 | 0.06 | 35.67 | 3.60 |
| Massachusetts | NH 2+ races | 0.01 | 0.02 | 15.82 | 2.90 |
| Massachusetts | NH Native | 0.00 | 0.00 | 66.73 | 7.02 |
| Massachusetts | NH NHPI | 0.00 | 0.00 | 0.00 | 0.00 |
| Michigan | All races | 1.00 | 1.00 | 43.89 | 6.37 |
| Michigan | NH White | 0.84 | 0.73 | 31.60 | 3.94 |
| Michigan | Hispanic | 0.02 | 0.05 | 69.59 | 8.83 |
| Michigan | NH Black | 0.11 | 0.15 | 137.70 | 19.94 |
| Michigan | NH Asian | 0.02 | 0.03 | 30.74 | 1.85 |
| Michigan | NH 2+ races | 0.01 | 0.03 | 8.51 | 1.30 |
| Michigan | NH Native | 0.00 | 0.01 | 81.21 | 10.05 |
| Michigan | NH NHPI | 0.00 | 0.00 | NA | NA |
| Minnesota | All races | 1.00 | 1.00 | 24.04 | 3.45 |
| Minnesota | NH White | 0.92 | 0.76 | 20.49 | 2.16 |
| Minnesota | Hispanic | 0.01 | 0.08 | 60.41 | 6.03 |
| Minnesota | NH Black | 0.03 | 0.07 | 76.44 | 9.46 |
| Minnesota | NH Asian | 0.02 | 0.05 | 66.43 | 7.93 |
| Minnesota | NH 2+ races | 0.01 | 0.03 | 25.93 | 2.31 |
| Minnesota | NH Native | 0.01 | 0.01 | 67.44 | 21.21 |
| Minnesota | NH NHPI | 0.00 | 0.00 | NA | NA |
| Mississippi | All races | 1.00 | 1.00 | 72.40 | 13.57 |
| Mississippi | NH White | 0.68 | 0.51 | 51.25 | 7.82 |
| Mississippi | Hispanic | 0.01 | 0.04 | 50.96 | 14.30 |
| Mississippi | NH Black | 0.30 | 0.43 | 117.16 | 19.20 |
| Mississippi | NH Asian | 0.01 | 0.01 | 110.70 | 15.06 |
| Mississippi | NH 2+ races | 0.01 | 0.01 | 52.92 | 6.48 |
| Mississippi | NH Native | 0.00 | 0.00 | 312.33 | 237.19 |
| Mississippi | NH NHPI | 0.00 | 0.00 | NA | NA |
| Missouri | All races | 1.00 | 1.00 | 38.33 | 5.08 |
| Missouri | NH White | 0.88 | 0.78 | 34.02 | 4.08 |
| Missouri | Hispanic | 0.01 | 0.05 | 70.77 | 10.10 |
| Missouri | NH Black | 0.08 | 0.12 | 78.80 | 9.87 |
| Missouri | NH Asian | 0.01 | 0.02 | 40.53 | 4.21 |
| Missouri | NH 2+ races | 0.01 | 0.03 | 15.69 | 1.58 |
| Missouri | NH Native | 0.00 | 0.00 | 39.15 | 5.87 |
| Missouri | NH NHPI | 0.00 | 0.00 | 252.75 | 22.85 |
| Montana | All races | 1.00 | 1.00 | 32.72 | 7.56 |
| Montana | NH White | 0.92 | 0.84 | 22.11 | 3.52 |
| Montana | Hispanic | 0.03 | 0.04 | 49.17 | 12.85 |
| Montana | NH Black | 0.00 | 0.01 | 222.08 | 0.00 |
| Montana | NH Asian | 0.00 | 0.00 | 159.58 | 49.19 |
| Montana | NH 2+ races | 0.01 | 0.04 | 48.41 | 5.81 |
| Montana | NH Native | 0.03 | 0.06 | 289.65 | 48.07 |
| Montana | NH NHPI | 0.00 | 0.00 | NA | NA |
| Nebraska | All races | 1.00 | 1.00 | 35.03 | 6.81 |
| Nebraska | NH White | 0.91 | 0.76 | 31.00 | 4.87 |
| Nebraska | Hispanic | 0.04 | 0.11 | 99.06 | 13.49 |
| Nebraska | NH Black | 0.03 | 0.06 | 66.85 | 14.42 |
| Nebraska | NH Asian | 0.02 | 0.04 | 28.31 | 8.64 |
| Nebraska | NH 2+ races | 0.01 | 0.02 | 88.86 | 5.09 |
| Nebraska | NH Native | 0.01 | 0.01 | 60.77 | 36.55 |
| Nebraska | NH NHPI | 0.00 | 0.00 | NA | NA |
| Nevada | All races | 1.00 | 1.00 | 46.69 | 10.17 |
| Nevada | NH White | 0.68 | 0.44 | 31.62 | 4.93 |
| Nevada | Hispanic | 0.12 | 0.32 | 98.95 | 17.38 |
| Nevada | NH Black | 0.07 | 0.09 | 71.22 | 15.73 |
| Nevada | NH Asian | 0.10 | 0.10 | 68.32 | 9.31 |
| Nevada | NH 2+ races | 0.01 | 0.04 | 23.45 | 6.02 |
| Nevada | NH Native | 0.01 | 0.01 | 76.16 | 27.48 |
| Nevada | NH NHPI | 0.00 | 0.01 | 74.48 | 61.15 |
| New Hampshire | All races | 1.00 | 1.00 | 10.14 | 1.87 |
| New Hampshire | NH White | 0.97 | 0.91 | 9.11 | 1.47 |
| New Hampshire | Hispanic | 0.01 | 0.03 | 43.21 | 1.90 |
| New Hampshire | NH Black | 0.01 | 0.01 | 69.51 | 19.71 |
| New Hampshire | NH Asian | 0.01 | 0.02 | 51.67 | 10.29 |
| New Hampshire | NH 2+ races | 0.00 | 0.02 | 0.00 | 0.00 |
| New Hampshire | NH Native | 0.00 | 0.00 | 0.00 | 0.00 |
| New Hampshire | NH NHPI | 0.00 | 0.00 | NA | NA |
| New Jersey | All races | 1.00 | 1.00 | 72.77 | 15.04 |
| New Jersey | NH White | 0.69 | 0.57 | 49.75 | 7.89 |
| New Jersey | Hispanic | 0.12 | 0.22 | 148.74 | 25.56 |
| New Jersey | NH Black | 0.11 | 0.13 | 141.78 | 26.97 |
| New Jersey | NH Asian | 0.08 | 0.07 | 69.27 | 13.42 |
| New Jersey | NH 2+ races | 0.01 | 0.02 | 50.43 | 6.52 |
| New Jersey | NH Native | 0.00 | 0.00 | 90.19 | 0.00 |
| New Jersey | NH NHPI | 0.00 | 0.00 | 546.66 | NA |
| New Mexico | All races | 1.00 | 1.00 | 37.59 | 11.03 |
| New Mexico | NH White | 0.57 | 0.32 | 13.97 | 3.14 |
| New Mexico | Hispanic | 0.33 | 0.53 | 44.99 | 7.82 |
| New Mexico | NH Black | 0.02 | 0.03 | 25.23 | 13.15 |
| New Mexico | NH Asian | 0.01 | 0.02 | 29.52 | 1.87 |
| New Mexico | NH 2+ races | 0.01 | 0.02 | 40.56 | 25.18 |
| New Mexico | NH Native | 0.06 | 0.09 | 232.49 | 60.79 |
| New Mexico | NH NHPI | 0.00 | 0.00 | 0.00 | NA |
| New York | All races | 1.00 | 1.00 | 79.88 | 15.26 |
| New York | NH White | 0.67 | 0.52 | 44.78 | 6.33 |
| New York | Hispanic | 0.12 | 0.23 | 173.98 | 27.94 |
| New York | NH Black | 0.12 | 0.13 | 173.87 | 27.86 |
| New York | NH Asian | 0.07 | 0.10 | 97.62 | 19.38 |
| New York | NH 2+ races | 0.01 | 0.02 | 15.20 | 2.97 |
| New York | NH Native | 0.00 | 0.00 | 43.38 | 27.37 |
| New York | NH NHPI | 0.00 | 0.00 | 1322.49 | 672.03 |
| North Carolina | All races | 1.00 | 1.00 | 13.53 | 2.32 |
| North Carolina | NH White | 0.76 | 0.60 | 8.89 | 1.28 |
| North Carolina | Hispanic | 0.02 | 0.09 | 53.04 | 5.60 |
| North Carolina | NH Black | 0.19 | 0.25 | 27.33 | 3.44 |
| North Carolina | NH Asian | 0.02 | 0.02 | 16.20 | 2.07 |
| North Carolina | NH 2+ races | 0.01 | 0.03 | 0.00 | 0.00 |
| North Carolina | NH Native | 0.01 | 0.01 | 23.51 | 2.48 |
| North Carolina | NH NHPI | 0.00 | 0.00 | 0.00 | 45.96 |
| North Dakota | All races | 1.00 | 1.00 | 56.44 | 10.65 |
| North Dakota | NH White | 0.94 | 0.86 | 47.79 | 7.16 |
| North Dakota | Hispanic | 0.01 | 0.06 | 160.62 | 15.08 |
| North Dakota | NH Black | 0.01 | 0.01 | 96.19 | 0.00 |
| North Dakota | NH Asian | 0.01 | 0.01 | 235.10 | 0.00 |
| North Dakota | NH 2+ races | 0.01 | 0.02 | 0.00 | 3.83 |
| North Dakota | NH Native | 0.03 | 0.03 | 298.02 | 57.18 |
| North Dakota | NH NHPI | 0.00 | 0.00 | 0.00 | NA |
| Ohio | All races | 1.00 | 1.00 | 29.34 | 3.62 |
| Ohio | NH White | 0.87 | 0.78 | 26.74 | 3.21 |
| Ohio | Hispanic | 0.01 | 0.05 | 37.53 | 4.02 |
| Ohio | NH Black | 0.09 | 0.13 | 54.05 | 5.28 |
| Ohio | NH Asian | 0.01 | 0.02 | 22.72 | 3.56 |
| Ohio | NH 2+ races | 0.01 | 0.03 | 11.36 | 2.15 |
| Ohio | NH Native | 0.00 | 0.00 | 41.93 | 25.18 |
| Ohio | NH NHPI | 0.00 | 0.00 | 0.00 | 0.00 |
| Oklahoma | All races | 1.00 | 1.00 | 40.42 | 5.48 |
| Oklahoma | NH White | 0.81 | 0.62 | 33.29 | 4.28 |
| Oklahoma | Hispanic | 0.03 | 0.13 | 102.06 | 7.88 |
| Oklahoma | NH Black | 0.05 | 0.07 | 56.51 | 7.16 |
| Oklahoma | NH Asian | 0.02 | 0.02 | 40.75 | 7.02 |
| Oklahoma | NH 2+ races | 0.04 | 0.07 | 29.00 | 1.30 |
| Oklahoma | NH Native | 0.05 | 0.09 | 105.05 | 12.59 |
| Oklahoma | NH NHPI | 0.00 | 0.00 | 626.61 | 149.86 |
| Oregon | All races | 1.00 | 1.00 | 7.69 | 2.08 |
| Oregon | NH White | 0.89 | 0.72 | 6.15 | 1.11 |
| Oregon | Hispanic | 0.04 | 0.15 | 28.16 | 4.73 |
| Oregon | NH Black | 0.01 | 0.02 | 19.22 | 8.50 |
| Oregon | NH Asian | 0.03 | 0.05 | 13.83 | 2.97 |
| Oregon | NH 2+ races | 0.01 | 0.04 | 0.00 | 1.71 |
| Oregon | NH Native | 0.01 | 0.01 | 18.37 | 12.31 |
| Oregon | NH NHPI | 0.00 | 0.00 | 102.99 | 16.21 |
| Pennsylvania | All races | 1.00 | 1.00 | 34.02 | 4.84 |
| Pennsylvania | NH White | 0.87 | 0.74 | 26.93 | 3.48 |
| Pennsylvania | Hispanic | 0.03 | 0.08 | 82.59 | 7.45 |
| Pennsylvania | NH Black | 0.07 | 0.11 | 98.09 | 12.50 |
| Pennsylvania | NH Asian | 0.02 | 0.04 | 44.22 | 4.98 |
| Pennsylvania | NH 2+ races | 0.01 | 0.03 | 8.77 | 1.45 |
| Pennsylvania | NH Native | 0.00 | 0.00 | 92.46 | 0.00 |
| Pennsylvania | NH NHPI | 0.00 | 0.00 | 0.00 | NA |
| Rhode Island | All races | 1.00 | 1.00 | 38.78 | 5.23 |
| Rhode Island | NH White | 0.86 | 0.75 | 33.00 | 3.45 |
| Rhode Island | Hispanic | 0.07 | 0.13 | 75.62 | 3.48 |
| Rhode Island | NH Black | 0.04 | 0.05 | 79.75 | 9.13 |
| Rhode Island | NH Asian | 0.02 | 0.04 | 34.05 | 13.15 |
| Rhode Island | NH 2+ races | 0.01 | 0.03 | 68.51 | 2.93 |
| Rhode Island | NH Native | 0.00 | 0.00 | 273.81 | NA |
| Rhode Island | NH NHPI | 0.00 | 0.00 | NA | NA |
| South Carolina | All races | 1.00 | 1.00 | 37.02 | 6.09 |
| South Carolina | NH White | 0.74 | 0.61 | 26.72 | 3.75 |
| South Carolina | Hispanic | 0.02 | 0.05 | 49.61 | 9.01 |
| South Carolina | NH Black | 0.22 | 0.28 | 72.37 | 10.19 |
| South Carolina | NH Asian | 0.01 | 0.02 | 13.87 | 7.55 |
| South Carolina | NH 2+ races | 0.01 | 0.03 | 27.35 | 0.00 |
| South Carolina | NH Native | 0.00 | 0.00 | 44.23 | 6.58 |
| South Carolina | NH NHPI | 0.00 | 0.00 | 0.00 | 4.62 |
| South Dakota | All races | 1.00 | 1.00 | 52.60 | 10.19 |
| South Dakota | NH White | 0.93 | 0.80 | 41.77 | 5.81 |
| South Dakota | Hispanic | 0.01 | 0.06 | 117.05 | 25.60 |
| South Dakota | NH Black | 0.01 | 0.03 | 151.87 | 19.41 |
| South Dakota | NH Asian | 0.00 | 0.02 | 188.29 | 57.16 |
| South Dakota | NH 2+ races | 0.00 | 0.02 | 223.12 | 33.89 |
| South Dakota | NH Native | 0.05 | 0.08 | 203.48 | 21.30 |
| South Dakota | NH NHPI | 0.00 | 0.00 | 0.00 | NA |
| Tennessee | All races | 1.00 | 1.00 | 43.00 | 6.51 |
| Tennessee | NH White | 0.84 | 0.72 | 37.51 | 4.73 |
| Tennessee | Hispanic | 0.01 | 0.06 | 130.53 | 14.27 |
| Tennessee | NH Black | 0.13 | 0.18 | 75.40 | 11.09 |
| Tennessee | NH Asian | 0.01 | 0.02 | 29.43 | 5.42 |
| Tennessee | NH 2+ races | 0.01 | 0.02 | 15.99 | 1.96 |
| Tennessee | NH Native | 0.00 | 0.00 | 118.65 | 26.92 |
| Tennessee | NH NHPI | 0.00 | 0.00 | 0.00 | 381.18 |
| Texas | All races | 1.00 | 1.00 | 58.35 | 9.73 |
| Texas | NH White | 0.60 | 0.40 | 31.23 | 4.79 |
| Texas | Hispanic | 0.25 | 0.42 | 129.46 | 15.79 |
| Texas | NH Black | 0.10 | 0.12 | 63.09 | 8.62 |
| Texas | NH Asian | 0.04 | 0.04 | 26.83 | 3.46 |
| Texas | NH 2+ races | 0.01 | 0.02 | 5.94 | 0.75 |
| Texas | NH Native | 0.00 | 0.00 | 30.63 | 12.43 |
| Texas | NH NHPI | 0.00 | 0.00 | 88.77 | 36.85 |
| Utah | All races | 1.00 | 1.00 | 25.38 | 4.01 |
| Utah | NH White | 0.89 | 0.75 | 17.85 | 2.35 |
| Utah | Hispanic | 0.07 | 0.15 | 77.22 | 8.36 |
| Utah | NH Black | 0.00 | 0.01 | 89.15 | 2.39 |
| Utah | NH Asian | 0.02 | 0.04 | 55.58 | 4.22 |
| Utah | NH 2+ races | 0.01 | 0.03 | 79.53 | 4.61 |
| Utah | NH Native | 0.01 | 0.01 | 202.46 | 28.45 |
| Utah | NH NHPI | 0.00 | 0.00 | 196.52 | 27.78 |
| Vermont | All races | 1.00 | 1.00 | 3.07 | 1.19 |
| Vermont | NH White | 0.96 | 0.91 | 3.20 | 1.03 |
| Vermont | Hispanic | 0.00 | 0.04 | 0.00 | 0.00 |
| Vermont | NH Black | 0.01 | 0.01 | 0.00 | 0.00 |
| Vermont | NH Asian | 0.02 | 0.02 | 0.00 | 0.00 |
| Vermont | NH 2+ races | 0.01 | 0.02 | 0.00 | 0.00 |
| Vermont | NH Native | 0.00 | 0.00 | 0.00 | 0.00 |
| Vermont | NH NHPI | 0.00 | 0.00 | 0.00 | NA |
| Virginia | All races | 1.00 | 1.00 | 22.90 | 3.47 |
| Virginia | NH White | 0.73 | 0.59 | 17.01 | 1.99 |
| Virginia | Hispanic | 0.03 | 0.11 | 58.31 | 7.54 |
| Virginia | NH Black | 0.17 | 0.20 | 42.70 | 6.25 |
| Virginia | NH Asian | 0.05 | 0.06 | 16.53 | 2.81 |
| Virginia | NH 2+ races | 0.01 | 0.03 | 16.34 | 0.38 |
| Virginia | NH Native | 0.00 | 0.00 | 41.86 | 20.31 |
| Virginia | NH NHPI | 0.00 | 0.00 | 0.00 | 0.00 |
| Washington | All races | 1.00 | 1.00 | 14.99 | 2.72 |
| Washington | NH White | 0.84 | 0.66 | 11.67 | 1.48 |
| Washington | Hispanic | 0.04 | 0.14 | 62.46 | 5.19 |
| Washington | NH Black | 0.03 | 0.05 | 28.11 | 4.94 |
| Washington | NH Asian | 0.07 | 0.09 | 16.61 | 2.97 |
| Washington | NH 2+ races | 0.01 | 0.05 | 8.61 | 1.89 |
| Washington | NH Native | 0.01 | 0.01 | 55.94 | 13.99 |
| Washington | NH NHPI | 0.00 | 0.01 | 72.94 | 22.64 |
| West Virginia | All races | 1.00 | 1.00 | 20.16 | 2.56 |
| West Virginia | NH White | 0.96 | 0.92 | 19.33 | 2.33 |
| West Virginia | Hispanic | 0.00 | 0.01 | 144.36 | 0.00 |
| West Virginia | NH Black | 0.03 | 0.04 | 48.03 | 5.65 |
| West Virginia | NH Asian | 0.01 | 0.01 | 0.00 | 3.29 |
| West Virginia | NH 2+ races | 0.01 | 0.01 | 0.00 | 0.00 |
| West Virginia | NH Native | 0.00 | 0.00 | 0.00 | 85.06 |
| West Virginia | NH NHPI | 0.00 | 0.00 | NA | NA |
| Wisconsin | All races | 1.00 | 1.00 | 31.37 | 4.47 |
| Wisconsin | NH White | 0.92 | 0.80 | 26.35 | 3.22 |
| Wisconsin | Hispanic | 0.02 | 0.08 | 123.32 | 11.23 |
| Wisconsin | NH Black | 0.03 | 0.06 | 83.00 | 8.63 |
| Wisconsin | NH Asian | 0.01 | 0.03 | 61.85 | 7.65 |
| Wisconsin | NH 2+ races | 0.01 | 0.02 | 23.54 | 0.00 |
| Wisconsin | NH Native | 0.01 | 0.01 | 110.99 | 20.34 |
| Wisconsin | NH NHPI | 0.00 | 0.00 | 1077.56 | 57.82 |
| Wyoming | All races | 1.00 | 1.00 | 20.61 | 5.39 |
| Wyoming | NH White | 0.92 | 0.81 | 16.04 | 2.58 |
| Wyoming | Hispanic | 0.04 | 0.13 | 35.59 | 5.81 |
| Wyoming | NH Black | 0.01 | 0.01 | 0.00 | 0.00 |
| Wyoming | NH Asian | 0.00 | 0.00 | 1123.29 | 93.15 |
| Wyoming | NH 2+ races | 0.01 | 0.03 | 0.00 | 2.82 |
| Wyoming | NH Native | 0.03 | 0.02 | 120.96 | 77.40 |
| Wyoming | NH NHPI | 0.00 | 0.00 | 0.00 | NA |
