## Supplementary material for "Fairness and efficiency considerations in COVID-19 vaccine allocation strategies: a case study comparing front-line workers and 65-74 year olds in the United States": 2022-09-12-Vaccine_prioritization_Supplementary_Table4_with95CI.docx

**S4 Table. Number of lives saved by vaccinating 1,000 individuals, either allocating doses to 65-74 year olds (policy S), or to front line workers (policy F). The ratio** $\frac{\mathbf{LS}_{\mathbf{j}}}{\mathbf{LF}_{\mathbf{j}}}$ **shows the relative number of lives saved comparing the two policies. Results shown for each US state per race/ethnicity categories.**

"NH" stands for "Non-Hispanic", "2+ races" stands for "Two or more races", "Native" stands for "American Indian or Alaska Native", "NHPI" stands for "Native Hawaiian or Other Pacific Islander"

| **State** | **Race/ethnicity** | ${LS}_{j}$ | ${LF}_{j}$ | **Ratio L (95% interval)** |
| --- | --- | --- | --- | --- |
| Alabama | All races | 0.5057 | 0.0857 | 5.93 (4.95; 7.05) |
| Alabama | NH White | 0.3161 | 0.0352 | 9.06 (7.02; 11.47) |
| Alabama | Hispanic | 0.0073 | 0.0054 | 1.48 (0.33; 3.8) |
| Alabama | NH Black | 0.1779 | 0.0421 | 4.26 (3.22; 5.55) |
| Alabama | NH Asian | 0.0017 | 0.0016 | NA |
| Alabama | NH 2+ races | <0.0001 | 0.0006 | NA |
| Alabama | NH AIAN | 0.0017 | 0.0011 | NA |
| Alabama | NH NHPI | <0.0001 | <0.0001 | NA |
| Alaska | All races | 0.1293 | 0.0436 | 3.17 (1.2; 6.57) |
| Alaska | NH White | 0.0289 | 0.0047 | NA |
| Alaska | Hispanic | <0.0001 | 0.0014 | NA |
| Alaska | NH Black | 0.0133 | 0.0021 | NA |
| Alaska | NH Asian | 0.0132 | 0.0077 | NA |
| Alaska | NH 2+ races | 0.0132 | 0.0028 | NA |
| Alaska | NH AIAN | 0.0473 | 0.0088 | 6.83 (1.04; 21.6) |
| Alaska | NH NHPI | 0.0132 | 0.0100 | NA |
| Arizona | All races | 0.4302 | 0.1032 | 4.18 (3.6; 4.83) |
| Arizona | NH White | 0.1905 | 0.0220 | 8.76 (6.6; 11.37) |
| Arizona | Hispanic | 0.1615 | 0.0448 | 3.63 (2.88; 4.52) |
| Arizona | NH Black | 0.0151 | 0.0052 | 3.11 (1.29; 6.18) |
| Arizona | NH Asian | 0.0072 | 0.0022 | 3.91 (1; 10.66) |
| Arizona | NH 2+ races | 0.0041 | 0.0010 | 5.24 (0.76; 17.49) |
| Arizona | NH AIAN | 0.0509 | 0.0240 | 2.14 (1.43; 3.04) |
| Arizona | NH NHPI | 0.0012 | 0.0003 | NA |
| Arkansas | All races | 0.3943 | 0.0671 | 5.93 (4.58; 7.66) |
| Arkansas | NH White | 0.2837 | 0.0345 | 8.35 (5.9; 11.51) |
| Arkansas | Hispanic | 0.0187 | 0.0084 | 2.4 (0.74; 5.38) |
| Arkansas | NH Black | 0.0713 | 0.0143 | 5.19 (2.81; 8.8) |
| Arkansas | NH Asian | 0.0056 | 0.0022 | NA |
| Arkansas | NH 2+ races | 0.0028 | 0.0014 | NA |
| Arkansas | NH AIAN | 0.0027 | 0.0005 | NA |
| Arkansas | NH NHPI | 0.0083 | 0.0028 | NA |
| California | All races | 0.2820 | 0.0557 | 5.07 (4.65; 5.5) |
| California | NH White | 0.0730 | 0.0078 | 9.43 (7.77; 11.39) |
| California | Hispanic | 0.1540 | 0.0385 | 4 (3.59; 4.44) |
| California | NH Black | 0.0214 | 0.0037 | 5.9 (4.3; 7.91) |
| California | NH Asian | 0.0280 | 0.0041 | 6.97 (5.17; 9.19) |
| California | NH 2+ races | 0.0020 | 0.0004 | 6.19 (1.88; 15.18) |
| California | NH AIAN | 0.0019 | 0.0003 | 9.2 (2.16; 27.24) |
| California | NH NHPI | 0.0018 | 0.0007 | 2.91 (0.87; 6.72) |
| Colorado | All races | 0.2357 | 0.0375 | 6.34 (4.9; 8.04) |
| Colorado | NH White | 0.1344 | 0.0124 | 11.1 (7.4; 15.86) |
| Colorado | Hispanic | 0.0707 | 0.0187 | 3.85 (2.48; 5.71) |
| Colorado | NH Black | 0.0149 | 0.0033 | 5.01 (1.64; 11.5) |
| Colorado | NH Asian | 0.0097 | 0.0018 | 6.94 (1.54; 20.08) |
| Colorado | NH 2+ races | 0.0014 | 0.0006 | NA |
| Colorado | NH AIAN | 0.0037 | 0.0014 | 4.4 (0.28; 18.13) |
| Colorado | NH NHPI | 0.0014 | 0.0003 | NA |
| Connecticut | All races | 0.3676 | 0.0514 | 7.22 (5.57; 9.25) |
| Connecticut | NH White | 0.2343 | 0.0215 | 11.12 (7.67; 15.61) |
| Connecticut | Hispanic | 0.0513 | 0.0155 | 3.44 (1.79; 5.97) |
| Connecticut | NH Black | 0.0745 | 0.0133 | 5.79 (3.26; 9.35) |
| Connecticut | NH Asian | 0.0054 | 0.0020 | 3.62 (0.3; 13.79) |
| Connecticut | NH 2+ races | <0.0001 | <0.0001 | NA |
| Connecticut | NH AIAN | 0.0021 | <0.0001 | NA |
| Connecticut | NH NHPI | <0.0001 | <0.0001 | NA |
| Delaware | All races | 0.2051 | 0.0700 | 3.04 (1.71; 5.03) |
| Delaware | NH White | 0.1333 | 0.0287 | 5.07 (2.19; 9.97) |
| Delaware | Hispanic | 0.0151 | 0.0124 | NA |
| Delaware | NH Black | 0.0567 | 0.0199 | 3.16 (1; 7.36) |
| Delaware | NH Asian | <0.0001 | 0.0035 | NA |
| Delaware | NH 2+ races | <0.0001 | 0.0010 | NA |
| Delaware | NH AIAN | <0.0001 | 0.0045 | NA |
| Delaware | NH NHPI | <0.0001 | <0.0001 | NA |
| District of Columbia | All races | 0.8984 | 0.1434 | 6.37 (4.27; 9.11) |
| District of Columbia | NH White | 0.0736 | 0.0046 | 21.07 (3.67; 69.25) |
| District of Columbia | Hispanic | 0.1246 | 0.0498 | 2.69 (0.9; 5.93) |
| District of Columbia | NH Black | 0.6650 | 0.1135 | 6.03 (3.65; 9.42) |
| District of Columbia | NH Asian | 0.0185 | 0.0029 | NA |
| District of Columbia | NH 2+ races | 0.0184 | <0.0001 | NA |
| District of Columbia | NH AIAN | <0.0001 | 0.0028 | NA |
| District of Columbia | NH NHPI | <0.0001 | <0.0001 | NA |
| Florida | All races | 0.3342 | 0.0634 | 5.28 (4.78; 5.83) |
| Florida | NH White | 0.1608 | 0.0184 | 8.77 (7.46; 10.29) |
| Florida | Hispanic | 0.0888 | 0.0216 | 4.13 (3.45; 4.93) |
| Florida | NH Black | 0.0777 | 0.0229 | 3.41 (2.8; 4.12) |
| Florida | NH Asian | 0.0055 | 0.0014 | 4.09 (1.78; 8.15) |
| Florida | NH 2+ races | 0.0007 | 0.0005 | NA |
| Florida | NH AIAN | 0.0004 | 0.0004 | NA |
| Florida | NH NHPI | 0.0004 | 0.0001 | NA |
| Georgia | All races | 0.4250 | 0.0675 | 6.32 (5.49; 7.21) |
| Georgia | NH White | 0.2105 | 0.0217 | 9.75 (7.8; 12.07) |
| Georgia | Hispanic | 0.0257 | 0.0090 | 2.91 (1.74; 4.46) |
| Georgia | NH Black | 0.1778 | 0.0341 | 5.24 (4.26; 6.37) |
| Georgia | NH Asian | 0.0088 | 0.0014 | 7.58 (2.37; 18.69) |
| Georgia | NH 2+ races | 0.0009 | 0.0004 | NA |
| Georgia | NH AIAN | <0.0001 | 0.0004 | NA |
| Georgia | NH NHPI | 0.0009 | <0.0001 | NA |
| Hawaii | All races | 0.0934 | 0.0346 | 2.83 (1.35; 5.16) |
| Hawaii | NH White | 0.0058 | 0.0025 | NA |
| Hawaii | Hispanic | 0.0058 | 0.0062 | NA |
| Hawaii | NH Black | 0.0058 | 0.0010 | NA |
| Hawaii | NH Asian | 0.0397 | 0.0118 | 3.95 (1.07; 10.25) |
| Hawaii | NH 2+ races | 0.0059 | 0.0038 | NA |
| Hawaii | NH AIAN | <0.0001 | <0.0001 | NA |
| Hawaii | NH NHPI | 0.0304 | 0.0109 | 3.27 (0.78; 8.82) |
| Idaho | All races | 0.2638 | 0.0411 | 6.59 (4.2; 9.97) |
| Idaho | NH White | 0.2061 | 0.0229 | 9.48 (5.24; 16.04) |
| Idaho | Hispanic | 0.0402 | 0.0106 | 4.28 (1.32; 10.21) |
| Idaho | NH Black | 0.0045 | 0.0014 | NA |
| Idaho | NH Asian | 0.0045 | 0.0029 | NA |
| Idaho | NH 2+ races | 0.0046 | 0.0013 | NA |
| Idaho | NH AIAN | 0.0045 | 0.0054 | NA |
| Idaho | NH NHPI | <0.0001 | <0.0001 | NA |
| Illinois | All races | 0.4322 | 0.0709 | 6.11 (5.39; 6.88) |
| Illinois | NH White | 0.2206 | 0.0193 | 11.51 (9.37; 14.04) |
| Illinois | Hispanic | 0.0858 | 0.0312 | 2.77 (2.17; 3.49) |
| Illinois | NH Black | 0.1056 | 0.0165 | 6.45 (4.99; 8.15) |
| Illinois | NH Asian | 0.0178 | 0.0028 | 6.69 (3.46; 11.94) |
| Illinois | NH 2+ races | 0.0007 | 0.0007 | NA |
| Illinois | NH AIAN | 0.0007 | 0.0002 | NA |
| Illinois | NH NHPI | 0.0007 | <0.0001 | NA |
| Indiana | All races | 0.3845 | 0.0495 | 7.8 (6.45; 9.38) |
| Indiana | NH White | 0.3038 | 0.0308 | 9.95 (7.91; 12.41) |
| Indiana | Hispanic | 0.0201 | 0.0056 | 3.74 (1.73; 6.82) |
| Indiana | NH Black | 0.0543 | 0.0102 | 5.44 (3.36; 8.23) |
| Indiana | NH Asian | 0.0028 | 0.0007 | NA |
| Indiana | NH 2+ races | 0.0012 | 0.0006 | NA |
| Indiana | NH AIAN | 0.0012 | 0.0006 | NA |
| Indiana | NH NHPI | 0.0012 | NA | NA |
| Iowa | All races | 0.3411 | 0.0598 | 5.77 (4.35; 7.48) |
| Iowa | NH White | 0.3000 | 0.0404 | 7.54 (5.49; 10.13) |
| Iowa | Hispanic | 0.0130 | 0.0059 | 2.47 (0.58; 6.38) |
| Iowa | NH Black | 0.0141 | 0.0082 | 1.96 (0.45; 5.1) |
| Iowa | NH Asian | 0.0074 | 0.0011 | 10.39 (0.89; 41.96) |
| Iowa | NH 2+ races | 0.0024 | 0.0009 | NA |
| Iowa | NH AIAN | 0.0023 | 0.0019 | NA |
| Iowa | NH NHPI | 0.0024 | <0.0001 | NA |
| Kansas | All races | 0.3078 | 0.0541 | 5.76 (4.23; 7.72) |
| Kansas | NH White | 0.2292 | 0.0269 | 8.72 (5.85; 12.53) |
| Kansas | Hispanic | 0.0370 | 0.0130 | 3.02 (1.25; 5.95) |
| Kansas | NH Black | 0.0239 | 0.0069 | 3.91 (1.18; 9.36) |
| Kansas | NH Asian | 0.0057 | 0.0022 | NA |
| Kansas | NH 2+ races | 0.0028 | 0.0011 | NA |
| Kansas | NH AIAN | 0.0068 | 0.0035 | NA |
| Kansas | NH NHPI | 0.0029 | <0.0001 | NA |
| Kentucky | All races | 0.2874 | 0.0368 | 7.9 (6.03; 10.17) |
| Kentucky | NH White | 0.2381 | 0.0254 | 9.49 (7.01; 12.65) |
| Kentucky | Hispanic | 0.0052 | 0.0051 | 1.13 (0.16; 3.38) |
| Kentucky | NH Black | 0.0405 | 0.0052 | 8.39 (3.97; 15.79) |
| Kentucky | NH Asian | 0.0018 | 0.0015 | NA |
| Kentucky | NH 2+ races | <0.0001 | <0.0001 | NA |
| Kentucky | NH AIAN | 0.0019 | <0.0001 | NA |
| Kentucky | NH NHPI | <0.0001 | <0.0001 | NA |
| Louisiana | All races | 0.5949 | 0.0994 | 6.01 (5.04; 7.1) |
| Louisiana | NH White | 0.2832 | 0.0282 | 10.18 (7.65; 13.36) |
| Louisiana | Hispanic | 0.0158 | 0.0066 | 2.6 (0.89; 5.77) |
| Louisiana | NH Black | 0.2906 | 0.0649 | 4.51 (3.56; 5.64) |
| Louisiana | NH Asian | 0.0020 | 0.0023 | NA |
| Louisiana | NH 2+ races | 0.0020 | <0.0001 | NA |
| Louisiana | NH AIAN | 0.0020 | 0.0012 | NA |
| Louisiana | NH NHPI | <0.0001 | 0.0006 | NA |
| Maine | All races | 0.0607 | 0.0116 | 5.94 (2.09; 13.57) |
| Maine | NH White | 0.0492 | 0.0076 | 7.84 (2.26; 20.57) |
| Maine | Hispanic | 0.0038 | <0.0001 | NA |
| Maine | NH Black | 0.0038 | 0.0027 | NA |
| Maine | NH Asian | <0.0001 | <0.0001 | NA |
| Maine | NH 2+ races | 0.0038 | <0.0001 | NA |
| Maine | NH AIAN | <0.0001 | <0.0001 | NA |
| Maine | NH NHPI | <0.0001 | <0.0001 | NA |
| Maryland | All races | 0.3552 | 0.0625 | 5.71 (4.71; 6.85) |
| Maryland | NH White | 0.1374 | 0.0128 | 10.94 (7.52; 15.55) |
| Maryland | Hispanic | 0.0348 | 0.0196 | 1.8 (1.04; 2.81) |
| Maryland | NH Black | 0.1652 | 0.0268 | 6.23 (4.67; 8.15) |
| Maryland | NH Asian | 0.0145 | 0.0032 | 5.06 (1.69; 11.91) |
| Maryland | NH 2+ races | 0.0015 | 0.0005 | NA |
| Maryland | NH AIAN | 0.0014 | <0.0001 | NA |
| Maryland | NH NHPI | <0.0001 | <0.0001 | NA |
| Massachusetts | All races | 0.3276 | 0.0444 | 7.43 (6.05; 9.03) |
| Massachusetts | NH White | 0.2412 | 0.0233 | 10.48 (8.1; 13.41) |
| Massachusetts | Hispanic | 0.0344 | 0.0102 | 3.46 (1.96; 5.6) |
| Massachusetts | NH Black | 0.0359 | 0.0078 | 4.74 (2.66; 7.82) |
| Massachusetts | NH Asian | 0.0142 | 0.0022 | 7.65 (2.56; 18.36) |
| Massachusetts | NH 2+ races | 0.0011 | 0.0007 | NA |
| Massachusetts | NH AIAN | 0.0011 | <0.0001 | NA |
| Massachusetts | NH NHPI | <0.0001 | <0.0001 | NA |
| Michigan | All races | 0.4172 | 0.0606 | 6.9 (6; 7.89) |
| Michigan | NH White | 0.2507 | 0.0275 | 9.18 (7.54; 11.11) |
| Michigan | Hispanic | 0.0151 | 0.0044 | 3.56 (1.8; 6.27) |
| Michigan | NH Black | 0.1408 | 0.0279 | 5.09 (4.04; 6.33) |
| Michigan | NH Asian | 0.0059 | 0.0005 | 14.39 (3.08; 46.01) |
| Michigan | NH 2+ races | 0.0008 | 0.0004 | NA |
| Michigan | NH AIAN | 0.0027 | 0.0006 | 6.95 (0.78; 26.91) |
| Michigan | NH NHPI | 0.0008 | <0.0001 | NA |
| Minnesota | All races | 0.2281 | 0.0327 | 7.05 (5.37; 9.08) |
| Minnesota | NH White | 0.1797 | 0.0156 | 11.78 (8.3; 16.3) |
| Minnesota | Hispanic | 0.0070 | 0.0045 | 1.74 (0.38; 4.54) |
| Minnesota | NH Black | 0.0217 | 0.0061 | 3.76 (1.61; 7.35) |
| Minnesota | NH Asian | 0.0141 | 0.0037 | 4.21 (1.46; 9.42) |
| Minnesota | NH 2+ races | 0.0013 | 0.0007 | NA |
| Minnesota | NH AIAN | 0.0045 | 0.0025 | 2.26 (0.27; 7.66) |
| Minnesota | NH NHPI | <0.0001 | <0.0001 | NA |
| Mississippi | All races | 0.6873 | 0.1289 | 5.36 (4.4; 6.48) |
| Mississippi | NH White | 0.3299 | 0.0376 | 8.91 (6.45; 12.07) |
| Mississippi | Hispanic | 0.0030 | 0.0049 | NA |
| Mississippi | NH Black | 0.3327 | 0.0789 | 4.25 (3.22; 5.5) |
| Mississippi | NH Asian | 0.0072 | 0.0017 | 6.47 (0.52; 26.82) |
| Mississippi | NH 2+ races | 0.0031 | 0.0006 | NA |
| Mississippi | NH AIAN | 0.0115 | 0.0083 | 1.54 (0.31; 4.17) |
| Mississippi | NH NHPI | <0.0001 | NA | NA |
| Missouri | All races | 0.3642 | 0.0483 | 7.59 (6.19; 9.22) |
| Missouri | NH White | 0.2826 | 0.0300 | 9.51 (7.45; 11.95) |
| Missouri | Hispanic | 0.0099 | 0.0050 | 2.17 (0.7; 4.88) |
| Missouri | NH Black | 0.0619 | 0.0114 | 5.59 (3.49; 8.56) |
| Missouri | NH Asian | 0.0055 | 0.0008 | 10.87 (1.33; 42.68) |
| Missouri | NH 2+ races | 0.0013 | 0.0004 | NA |
| Missouri | NH AIAN | 0.0013 | 0.0002 | NA |
| Missouri | NH NHPI | 0.0013 | 0.0005 | NA |
| Montana | All races | 0.3107 | 0.0719 | 4.43 (2.82; 6.69) |
| Montana | NH White | 0.1928 | 0.0282 | 7.29 (3.75; 12.95) |
| Montana | Hispanic | 0.0138 | 0.0045 | NA |
| Montana | NH Black | 0.0064 | <0.0001 | NA |
| Montana | NH Asian | 0.0065 | 0.0012 | NA |
| Montana | NH 2+ races | 0.0063 | 0.0023 | NA |
| Montana | NH AIAN | 0.0849 | 0.0290 | 3.13 (1.31; 6.16) |
| Montana | NH NHPI | <0.0001 | NA | NA |
| Nebraska | All races | 0.3327 | 0.0647 | 5.23 (3.64; 7.31) |
| Nebraska | NH White | 0.2661 | 0.0351 | 7.8 (4.91; 11.78) |
| Nebraska | Hispanic | 0.0356 | 0.0147 | 2.61 (0.86; 5.77) |
| Nebraska | NH Black | 0.0173 | 0.0081 | 2.59 (0.44; 8.13) |
| Nebraska | NH Asian | 0.0044 | 0.0030 | NA |
| Nebraska | NH 2+ races | 0.0042 | 0.0011 | NA |
| Nebraska | NH AIAN | 0.0043 | 0.0027 | NA |
| Nebraska | NH NHPI | <0.0001 | <0.0001 | NA |
| Nevada | All races | 0.4435 | 0.0966 | 4.62 (3.64; 5.78) |
| Nevada | NH White | 0.2046 | 0.0206 | 10.23 (6.65; 15.07) |
| Nevada | Hispanic | 0.1126 | 0.0520 | 2.2 (1.43; 3.21) |
| Nevada | NH Black | 0.0481 | 0.0128 | 3.95 (1.9; 7.09) |
| Nevada | NH Asian | 0.0644 | 0.0092 | 7.6 (3.46; 14.66) |
| Nevada | NH 2+ races | 0.0030 | 0.0023 | NA |
| Nevada | NH AIAN | 0.0078 | 0.0016 | 6.97 (0.74; 26.34) |
| Nevada | NH NHPI | 0.0030 | 0.0051 | NA |
| New Hampshire | All races | 0.0962 | 0.0178 | 5.92 (2.53; 12) |
| New Hampshire | NH White | 0.0839 | 0.0127 | 7.61 (2.73; 17.44) |
| New Hampshire | Hispanic | 0.0041 | 0.0006 | NA |
| New Hampshire | NH Black | 0.0041 | 0.0017 | NA |
| New Hampshire | NH Asian | 0.0040 | 0.0024 | NA |
| New Hampshire | NH 2+ races | <0.0001 | <0.0001 | NA |
| New Hampshire | NH AIAN | <0.0001 | <0.0001 | NA |
| New Hampshire | NH NHPI | <0.0001 | <0.0001 | NA |
| New Jersey | All races | 0.6912 | 0.1429 | 4.85 (4.34; 5.39) |
| New Jersey | NH White | 0.3265 | 0.0423 | 7.75 (6.43; 9.31) |
| New Jersey | Hispanic | 0.1623 | 0.0530 | 3.08 (2.5; 3.75) |
| New Jersey | NH Black | 0.1457 | 0.0334 | 4.39 (3.44; 5.49) |
| New Jersey | NH Asian | 0.0510 | 0.0088 | 5.95 (3.83; 8.84) |
| New Jersey | NH 2+ races | 0.0035 | 0.0010 | 4.24 (0.61; 14.2) |
| New Jersey | NH AIAN | 0.0010 | <0.0001 | NA |
| New Jersey | NH NHPI | 0.0010 | <0.0001 | NA |
| New Mexico | All races | 0.3570 | 0.1046 | 3.44 (2.56; 4.53) |
| New Mexico | NH White | 0.0765 | 0.0094 | 8.91 (3.97; 17.94) |
| New Mexico | Hispanic | 0.1406 | 0.0393 | 3.67 (2.24; 5.67) |
| New Mexico | NH Black | 0.0037 | 0.0032 | NA |
| New Mexico | NH Asian | 0.0037 | 0.0003 | NA |
| New Mexico | NH 2+ races | 0.0037 | 0.0040 | NA |
| New Mexico | NH AIAN | 0.1289 | 0.0546 | 2.41 (1.48; 3.64) |
| New Mexico | NH NHPI | <0.0001 | <0.0001 | NA |
| New York | All races | 0.7589 | 0.1450 | 5.24 (4.87; 5.63) |
| New York | NH White | 0.2852 | 0.0315 | 9.08 (7.94; 10.37) |
| New York | Hispanic | 0.1998 | 0.0607 | 3.3 (2.9; 3.74) |
| New York | NH Black | 0.2030 | 0.0331 | 6.15 (5.33; 7.04) |
| New York | NH Asian | 0.0679 | 0.0186 | 3.67 (2.85; 4.63) |
| New York | NH 2+ races | 0.0014 | 0.0005 | 3.29 (0.43; 11.1) |
| New York | NH AIAN | 0.0011 | 0.0005 | 2.93 (0.28; 10.73) |
| New York | NH NHPI | 0.0005 | 0.0007 | NA |
| North Carolina | All races | 0.1285 | 0.0220 | 5.88 (4.59; 7.41) |
| North Carolina | NH White | 0.0640 | 0.0073 | 9 (6.08; 12.92) |
| North Carolina | Hispanic | 0.0115 | 0.0050 | 2.4 (1.14; 4.3) |
| North Carolina | NH Black | 0.0484 | 0.0081 | 6.09 (4.07; 8.9) |
| North Carolina | NH Asian | 0.0024 | 0.0004 | 8.13 (0.79; 31.82) |
| North Carolina | NH 2+ races | <0.0001 | <0.0001 | NA |
| North Carolina | NH AIAN | 0.0022 | 0.0003 | NA |
| North Carolina | NH NHPI | <0.0001 | 0.0001 | NA |
| North Dakota | All races | 0.5362 | 0.1013 | 5.43 (3.36; 8.29) |
| North Dakota | NH White | 0.4263 | 0.0588 | 7.6 (4.23; 12.84) |
| North Dakota | Hispanic | 0.0119 | 0.0091 | NA |
| North Dakota | NH Black | 0.0118 | <0.0001 | NA |
| North Dakota | NH Asian | 0.0117 | <0.0001 | NA |
| North Dakota | NH 2+ races | <0.0001 | 0.0007 | NA |
| North Dakota | NH AIAN | 0.0739 | 0.0187 | 4.47 (1.19; 11.4) |
| North Dakota | NH NHPI | <0.0001 | <0.0001 | NA |
| Ohio | All races | 0.2786 | 0.0344 | 8.13 (6.86; 9.57) |
| Ohio | NH White | 0.2207 | 0.0236 | 9.4 (7.71; 11.37) |
| Ohio | Hispanic | 0.0048 | 0.0019 | 2.83 (0.85; 6.66) |
| Ohio | NH Black | 0.0487 | 0.0065 | 7.68 (5.1; 11.05) |
| Ohio | NH Asian | 0.0031 | 0.0007 | 5.97 (1.04; 18.9) |
| Ohio | NH 2+ races | 0.0006 | 0.0005 | NA |
| Ohio | NH AIAN | 0.0006 | 0.0005 | NA |
| Ohio | NH NHPI | <0.0001 | <0.0001 | NA |
| Oklahoma | All races | 0.3842 | 0.0520 | 7.46 (5.8; 9.45) |
| Oklahoma | NH White | 0.2566 | 0.0252 | 10.37 (7.39; 14.13) |
| Oklahoma | Hispanic | 0.0278 | 0.0096 | 3.08 (1.28; 6.06) |
| Oklahoma | NH Black | 0.0289 | 0.0049 | 6.58 (2.52; 14.31) |
| Oklahoma | NH Asian | 0.0060 | 0.0014 | 7.52 (0.53; 34.25) |
| Oklahoma | NH 2+ races | 0.0107 | 0.0009 | NA |
| Oklahoma | NH AIAN | 0.0520 | 0.0102 | 5.32 (2.73; 9.27) |
| Oklahoma | NH NHPI | 0.0023 | 0.0031 | NA |
| Oregon | All races | 0.0732 | 0.0198 | 3.78 (2.36; 5.69) |
| Oregon | NH White | 0.0519 | 0.0076 | 7.16 (3.78; 12.55) |
| Oregon | Hispanic | 0.0119 | 0.0067 | 1.89 (0.59; 4.36) |
| Oregon | NH Black | 0.0017 | 0.0014 | NA |
| Oregon | NH Asian | 0.0043 | 0.0014 | 5.16 (0.35; 22.13) |
| Oregon | NH 2+ races | <0.0001 | 0.0007 | NA |
| Oregon | NH AIAN | 0.0017 | 0.0014 | NA |
| Oregon | NH NHPI | 0.0017 | 0.0008 | NA |
| Pennsylvania | All races | 0.3231 | 0.0460 | 7.04 (6.1; 8.1) |
| Pennsylvania | NH White | 0.2223 | 0.0245 | 9.1 (7.59; 10.86) |
| Pennsylvania | Hispanic | 0.0213 | 0.0054 | 4.02 (2.42; 6.22) |
| Pennsylvania | NH Black | 0.0691 | 0.0135 | 5.16 (3.84; 6.71) |
| Pennsylvania | NH Asian | 0.0092 | 0.0018 | 5.48 (2.21; 11.71) |
| Pennsylvania | NH 2+ races | 0.0006 | 0.0004 | NA |
| Pennsylvania | NH AIAN | 0.0006 | <0.0001 | NA |
| Pennsylvania | NH NHPI | <0.0001 | <0.0001 | NA |
| Rhode Island | All races | 0.3680 | 0.0498 | 7.63 (4.68; 11.9) |
| Rhode Island | NH White | 0.2698 | 0.0248 | 11.67 (6.02; 20.95) |
| Rhode Island | Hispanic | 0.0503 | 0.0041 | 15.93 (3.42; 48.42) |
| Rhode Island | NH Black | 0.0290 | 0.0043 | 10.51 (1.22; 40.98) |
| Rhode Island | NH Asian | 0.0066 | 0.0047 | NA |
| Rhode Island | NH 2+ races | 0.0066 | 0.0008 | NA |
| Rhode Island | NH AIAN | 0.0066 | <0.0001 | NA |
| Rhode Island | NH NHPI | <0.0001 | <0.0001 | NA |
| South Carolina | All races | 0.3516 | 0.0579 | 6.11 (4.96; 7.47) |
| South Carolina | NH White | 0.1890 | 0.0218 | 8.81 (6.44; 11.85) |
| South Carolina | Hispanic | 0.0076 | 0.0047 | 1.73 (0.46; 4.07) |
| South Carolina | NH Black | 0.1507 | 0.0272 | 5.6 (4.1; 7.48) |
| South Carolina | NH Asian | 0.0015 | 0.0014 | NA |
| South Carolina | NH 2+ races | 0.0015 | <0.0001 | NA |
| South Carolina | NH AIAN | 0.0015 | 0.0001 | NA |
| South Carolina | NH NHPI | <0.0001 | 0.0002 | NA |
| South Dakota | All races | 0.4994 | 0.0968 | 5.3 (3.36; 8.06) |
| South Dakota | NH White | 0.3662 | 0.0439 | 8.8 (4.85; 15.02) |
| South Dakota | Hispanic | 0.0088 | 0.0139 | NA |
| South Dakota | NH Black | 0.0088 | 0.0048 | NA |
| South Dakota | NH Asian | 0.0086 | 0.0130 | NA |
| South Dakota | NH 2+ races | 0.0089 | 0.0063 | NA |
| South Dakota | NH AIAN | 0.0969 | 0.0154 | 7.3 (2.33; 17.47) |
| South Dakota | NH NHPI | <0.0001 | <0.0001 | NA |
| Tennessee | All races | 0.4084 | 0.0619 | 6.63 (5.56; 7.85) |
| Tennessee | NH White | 0.3007 | 0.0324 | 9.36 (7.47; 11.57) |
| Tennessee | Hispanic | 0.0125 | 0.0077 | 1.7 (0.69; 3.35) |
| Tennessee | NH Black | 0.0895 | 0.0189 | 4.79 (3.37; 6.5) |
| Tennessee | NH Asian | 0.0033 | 0.0010 | 4.6 (0.47; 16.83) |
| Tennessee | NH 2+ races | 0.0013 | 0.0004 | NA |
| Tennessee | NH AIAN | 0.0013 | 0.0006 | NA |
| Tennessee | NH NHPI | <0.0001 | 0.0028 | NA |
| Texas | All races | 0.5543 | 0.0924 | 6 (5.56; 6.46) |
| Texas | NH White | 0.1771 | 0.0182 | 9.77 (8.41; 11.3) |
| Texas | Hispanic | 0.3026 | 0.0625 | 4.84 (4.39; 5.33) |
| Texas | NH Black | 0.0620 | 0.0099 | 6.28 (5.01; 7.74) |
| Texas | NH Asian | 0.0109 | 0.0013 | 8.87 (4.76; 15.35) |
| Texas | NH 2+ races | 0.0004 | 0.0001 | NA |
| Texas | NH AIAN | 0.0009 | 0.0004 | 2.48 (0.25; 8.65) |
| Texas | NH NHPI | 0.0004 | 0.0004 | NA |
| Utah | All races | 0.2417 | 0.0380 | 6.46 (4.4; 9.2) |
| Utah | NH White | 0.1516 | 0.0168 | 9.35 (5.46; 15.04) |
| Utah | Hispanic | 0.0503 | 0.0123 | 4.36 (1.85; 8.61) |
| Utah | NH Black | 0.0038 | 0.0003 | NA |
| Utah | NH Asian | 0.0083 | 0.0015 | NA |
| Utah | NH 2+ races | 0.0038 | 0.0012 | NA |
| Utah | NH AIAN | 0.0151 | 0.0029 | 6.66 (1.07; 21.8) |
| Utah | NH NHPI | 0.0082 | 0.0013 | NA |
| Vermont | All races | 0.0293 | 0.0112 | 3.49 (0.35; 13.13) |
| Vermont | NH White | 0.0289 | 0.0089 | 4.94 (0.41; 18.99) |
| Vermont | Hispanic | <0.0001 | <0.0001 | NA |
| Vermont | NH Black | <0.0001 | <0.0001 | NA |
| Vermont | NH Asian | <0.0001 | <0.0001 | NA |
| Vermont | NH 2+ races | <0.0001 | <0.0001 | NA |
| Vermont | NH AIAN | <0.0001 | <0.0001 | NA |
| Vermont | NH NHPI | <0.0001 | <0.0001 | NA |
| Virginia | All races | 0.2177 | 0.0329 | 6.66 (5.35; 8.19) |
| Virginia | NH White | 0.1181 | 0.0112 | 10.71 (7.65; 14.65) |
| Virginia | Hispanic | 0.0193 | 0.0078 | 2.55 (1.31; 4.35) |
| Virginia | NH Black | 0.0699 | 0.0121 | 5.86 (3.99; 8.33) |
| Virginia | NH Asian | 0.0083 | 0.0016 | 5.94 (1.68; 15.45) |
| Virginia | NH 2+ races | 0.0010 | 0.0001 | NA |
| Virginia | NH AIAN | 0.0010 | 0.0002 | NA |
| Virginia | NH NHPI | <0.0001 | <0.0001 | NA |
| Washington | All races | 0.1424 | 0.0258 | 5.57 (4.21; 7.22) |
| Washington | NH White | 0.0930 | 0.0093 | 10.25 (6.76; 15.01) |
| Washington | Hispanic | 0.0227 | 0.0069 | 3.43 (1.78; 5.84) |
| Washington | NH Black | 0.0071 | 0.0023 | 3.59 (0.93; 9.4) |
| Washington | NH Asian | 0.0112 | 0.0024 | 5.33 (1.73; 12.79) |
| Washington | NH 2+ races | 0.0011 | 0.0008 | NA |
| Washington | NH AIAN | 0.0049 | 0.0016 | 3.7 (0.73; 10.7) |
| Washington | NH NHPI | 0.0024 | 0.0014 | NA |
| West Virginia | All races | 0.1914 | 0.0243 | 8.22 (4.87; 13.2) |
| West Virginia | NH White | 0.1758 | 0.0204 | 9.09 (5.12; 15.37) |
| West Virginia | Hispanic | 0.0039 | <0.0001 | NA |
| West Virginia | NH Black | 0.0116 | 0.0024 | NA |
| West Virginia | NH Asian | <0.0001 | 0.0003 | NA |
| West Virginia | NH 2+ races | <0.0001 | <0.0001 | NA |
| West Virginia | NH AIAN | <0.0001 | 0.0025 | NA |
| West Virginia | NH NHPI | <0.0001 | <0.0001 | NA |
| Wisconsin | All races | 0.2982 | 0.0424 | 7.09 (5.61; 8.84) |
| Wisconsin | NH White | 0.2305 | 0.0244 | 9.58 (7.18; 12.59) |
| Wisconsin | Hispanic | 0.0245 | 0.0086 | 2.96 (1.45; 5.39) |
| Wisconsin | NH Black | 0.0268 | 0.0050 | 5.73 (2.66; 10.48) |
| Wisconsin | NH Asian | 0.0070 | 0.0023 | 3.71 (0.81; 10.33) |
| Wisconsin | NH 2+ races | 0.0014 | <0.0001 | NA |
| Wisconsin | NH AIAN | 0.0065 | 0.0018 | 4.72 (0.8; 15.16) |
| Wisconsin | NH NHPI | 0.0014 | 0.0008 | NA |
| Wyoming | All races | 0.1961 | 0.0512 | 4.1 (1.75; 8.11) |
| Wyoming | NH White | 0.1392 | 0.0198 | 8.32 (2.55; 21.32) |
| Wyoming | Hispanic | 0.0127 | 0.0071 | NA |
| Wyoming | NH Black | <0.0001 | <0.0001 | NA |
| Wyoming | NH Asian | 0.0127 | 0.0039 | NA |
| Wyoming | NH 2+ races | <0.0001 | 0.0007 | NA |
| Wyoming | NH AIAN | 0.0303 | 0.0171 | NA |
| Wyoming | NH NHPI | <0.0001 | <0.0001 | NA |
