## Supplementary material for "Fairness and efficiency considerations in COVID-19 vaccine allocation strategies: a case study comparing front-line workers and 65-74 year olds in the United States": 2022-09-12-Vaccine_prioritization_Supplementary_Table5_with95CI.docx

**S5 Table. Relative number of lives saved comparing a policy allocating vaccines to 65-74 year olds (policy S) to one allocating vaccines to front-line workers (policy F). Results shown for varying values of *R*, the increased risk of infection for front-line workers.** The ratios for an R value of 1·56 correspond to those shown in Table 3. "NH" stands for "Non-Hispanic", "2+ races" stands for "Two or more races", "Native" stands for "American Indian or Alaska Native", "NHPI" stands for "Native Hawaiian or Other Pacific Islander".

| **Race/ethnicity** | **Ratio L (95% uncertainty interval)** | | | | |
| --- | --- | --- | --- | --- | --- |
|  | ***R* = 1** | ***R* = 1·56** | ***R* = 2** | ***R* = 3** | ***R* = 4** |
| All races | 8·92 (8·69; 9·15) | 5·72 (5·57; 5·86) | 4·46 (4·35; 4·57) | 2·97 (2·9; 3·05) | 2·23 (2·17; 2·29) |
| Non-Hispanic White | 14·53 (13·95; 15·11) | 9·31 (8·94; 9·68) | 7·26 (6·97; 7·55) | 4·84 (4·65; 5·03) | 3·63 (3·49; 3·78) |
| Hispanic | 5·42 (5·16; 5·69) | 3·47 (3·31; 3·64) | 2·71 (2·58; 2·84) | 1·81 (1·72; 1·9) | 1·36 (1·29; 1·42) |
| Non-Hispanic Black | 7·93 (7·51; 8·35) | 5·09 (4·82; 5·36) | 3·97 (3·76; 4·18) | 2·64 (2·51; 2·79) | 1·98 (1·88; 2·09) |
| Non-Hispanic Asian | 7·73 (6·81; 8·72) | 4·95 (4·36; 5·6) | 3·86 (3·41; 4·34) | 2·58 (2·27; 2·91) | 1·93 (1·71; 2·18) |
| Non-Hispanic 2+ races | 4·24 (2·96; 5·82) | 2·71 (1·9; 3·77) | 2·11 (1·48; 2·91) | 1·41 (**0·98**; 1·95) | 1·06 (**0·73**; 1·46) |
| Non-Hispanic Native | 4·73 (3·91; 5·67) | 3·03 (2·5; 3·63) | 2·37 (1·96; 2·84) | 1·58 (1·31; 1·9) | 1·18 (**0·98**; 1·41) |
| Non-Hispanic NHPI | 2·4 (1·58; 3·47) | 1·54 (1·01; 2·24) | 1·2 (**0·79**; 1·74) | **0·8** (**0·52**; 1·16) | **0·6** (**0·39**; **0·87**) |
