## Supplementary material for "Fairness and efficiency considerations in COVID-19 vaccine allocation strategies: a case study comparing front-line workers and 65-74 year olds in the United States": 2022-09-12-Vaccine_prioritization_Supplementary_Table6_with95CI.docx

**S6 Table. Number of years of life saved by vaccinating 1,000 individuals, either allocating doses to 65-74 year olds (policy S), or to front line workers (policy F). The ratio** $\frac{\mathbf{YS}_{\mathbf{j}}}{\mathbf{YF}_{\mathbf{j}}}$ **shows the relative number of years of life saved comparing the two policies. Results shown for each US state per race/ethnicity categories.**

"NH" stands for "Non-Hispanic", "2+ races" stands for "Two or more races", "Native" stands for "American Indian or Alaska Native", "NHPI" stands for "Native Hawaiian or Other Pacific Islander"

| **State** | **Race** | ${YS}_{j}$ | ${YF}_{j}$ | **Ratio Y (95% interval)** |
| --- | --- | --- | --- | --- |
| Alabama | All races | 8.8031 | 2.7161 | 3.26 (2.7; 3.9) |
| Alabama | NH White | 5.5096 | 1.0688 | 5.22 (4; 6.71) |
| Alabama | Hispanic | 0.1281 | 0.1881 | 0.75 (0.17; 2.01) |
| Alabama | NH Black | 3.1033 | 1.3915 | 2.26 (1.66; 2.99) |
| Alabama | NH Asian | 0.0307 | 0.0531 | NA |
| Alabama | NH 2+ races | <0.0001 | 0.0187 | NA |
| Alabama | NH AIAN | 0.0307 | 0.0371 | NA |
| Alabama | NH NHPI | <0.0001 | <0.0001 | NA |
| Alaska | All races | 2.2736 | 1.5288 | 1.61 (0.6; 3.39) |
| Alaska | NH White | 0.4974 | 0.1649 | NA |
| Alaska | Hispanic | <0.0001 | 0.0388 | NA |
| Alaska | NH Black | 0.2304 | 0.0704 | NA |
| Alaska | NH Asian | 0.2291 | 0.2582 | NA |
| Alaska | NH 2+ races | 0.2313 | 0.0715 | NA |
| Alaska | NH AIAN | 0.8178 | 0.3450 | 3.11 (0.45; 10.38) |
| Alaska | NH NHPI | 0.2296 | 0.2583 | NA |
| Arizona | All races | 7.4948 | 3.3438 | 2.25 (1.93; 2.61) |
| Arizona | NH White | 3.3172 | 0.6496 | 5.18 (3.86; 6.84) |
| Arizona | Hispanic | 2.8138 | 1.4593 | 1.94 (1.51; 2.42) |
| Arizona | NH Black | 0.2638 | 0.1696 | 1.68 (0.68; 3.39) |
| Arizona | NH Asian | 0.1252 | 0.0685 | 2.15 (0.55; 5.84) |
| Arizona | NH 2+ races | 0.0709 | 0.0401 | 2.37 (0.33; 8.19) |
| Arizona | NH AIAN | 0.8862 | 0.8413 | 1.07 (0.7; 1.53) |
| Arizona | NH NHPI | 0.0207 | 0.0118 | NA |
| Arkansas | All races | 6.8496 | 2.1182 | 3.27 (2.48; 4.26) |
| Arkansas | NH White | 4.9434 | 1.0591 | 4.77 (3.28; 6.62) |
| Arkansas | Hispanic | 0.3260 | 0.2565 | 1.37 (0.41; 3.13) |
| Arkansas | NH Black | 1.2419 | 0.4452 | 2.92 (1.56; 5.01) |
| Arkansas | NH Asian | 0.0970 | 0.0588 | NA |
| Arkansas | NH 2+ races | 0.0477 | 0.0419 | NA |
| Arkansas | NH AIAN | 0.0477 | 0.0131 | NA |
| Arkansas | NH NHPI | 0.1439 | 0.1580 | NA |
| California | All races | 4.9162 | 1.7841 | 2.76 (2.52; 3.01) |
| California | NH White | 1.2733 | 0.2342 | 5.48 (4.44; 6.69) |
| California | Hispanic | 2.6841 | 1.2585 | 2.14 (1.91; 2.38) |
| California | NH Black | 0.3726 | 0.1180 | 3.2 (2.28; 4.35) |
| California | NH Asian | 0.4868 | 0.1235 | 3.99 (2.94; 5.26) |
| California | NH 2+ races | 0.0348 | 0.0126 | 3.21 (0.93; 8.06) |
| California | NH AIAN | 0.0329 | 0.0102 | 4.37 (0.92; 13.7) |
| California | NH NHPI | 0.0310 | 0.0221 | 1.55 (0.47; 3.59) |
| Colorado | All races | 4.1107 | 1.2099 | 3.43 (2.6; 4.43) |
| Colorado | NH White | 2.3432 | 0.3810 | 6.35 (4.06; 9.35) |
| Colorado | Hispanic | 1.2247 | 0.5826 | 2.15 (1.37; 3.16) |
| Colorado | NH Black | 0.2584 | 0.1213 | 2.51 (0.72; 6.07) |
| Colorado | NH Asian | 0.1677 | 0.0544 | 3.94 (0.84; 11.72) |
| Colorado | NH 2+ races | 0.0244 | 0.0205 | NA |
| Colorado | NH AIAN | 0.0649 | 0.0620 | 1.91 (0.11; 8.6) |
| Colorado | NH NHPI | 0.0248 | 0.0063 | NA |
| Connecticut | All races | 6.4060 | 1.5459 | 4.19 (3.17; 5.44) |
| Connecticut | NH White | 4.0891 | 0.6069 | 6.9 (4.7; 9.86) |
| Connecticut | Hispanic | 0.8988 | 0.5110 | 1.84 (0.92; 3.25) |
| Connecticut | NH Black | 1.2971 | 0.3882 | 3.48 (1.94; 5.85) |
| Connecticut | NH Asian | 0.0933 | 0.0644 | 2.01 (0.16; 8.06) |
| Connecticut | NH 2+ races | <0.0001 | <0.0001 | NA |
| Connecticut | NH AIAN | 0.0367 | <0.0001 | NA |
| Connecticut | NH NHPI | <0.0001 | <0.0001 | NA |
| Delaware | All races | 3.5748 | 2.6130 | 1.44 (0.75; 2.43) |
| Delaware | NH White | 2.3180 | 1.0487 | 2.49 (0.97; 5.17) |
| Delaware | Hispanic | 0.2667 | 0.5427 | NA |
| Delaware | NH Black | 0.9841 | 0.7428 | 1.52 (0.43; 3.67) |
| Delaware | NH Asian | <0.0001 | 0.0890 | NA |
| Delaware | NH 2+ races | <0.0001 | 0.0248 | NA |
| Delaware | NH AIAN | <0.0001 | 0.1130 | NA |
| Delaware | NH NHPI | <0.0001 | <0.0001 | NA |
| District of Columbia | All races | 15.6514 | 4.5162 | 3.54 (2.31; 5.11) |
| District of Columbia | NH White | 1.2796 | 0.1451 | 11.82 (1.93; 38.98) |
| District of Columbia | Hispanic | 2.1676 | 1.7598 | 1.35 (0.44; 3.07) |
| District of Columbia | NH Black | 11.6152 | 3.4654 | 3.47 (2.04; 5.52) |
| District of Columbia | NH Asian | 0.3150 | 0.1222 | NA |
| District of Columbia | NH 2+ races | 0.3155 | <0.0001 | NA |
| District of Columbia | NH AIAN | <0.0001 | 0.0704 | NA |
| District of Columbia | NH NHPI | <0.0001 | <0.0001 | NA |
| Florida | All races | 5.8238 | 1.9766 | 2.95 (2.66; 3.27) |
| Florida | NH White | 2.8003 | 0.5398 | 5.22 (4.38; 6.2) |
| Florida | Hispanic | 1.5485 | 0.6653 | 2.34 (1.94; 2.79) |
| Florida | NH Black | 1.3521 | 0.7241 | 1.88 (1.53; 2.28) |
| Florida | NH Asian | 0.0956 | 0.0480 | 2.16 (0.94; 4.28) |
| Florida | NH 2+ races | 0.0132 | 0.0191 | NA |
| Florida | NH AIAN | 0.0066 | 0.0197 | NA |
| Florida | NH NHPI | 0.0066 | 0.0037 | NA |
| Georgia | All races | 7.4015 | 2.1317 | 3.48 (3.01; 4.01) |
| Georgia | NH White | 3.6708 | 0.6403 | 5.78 (4.58; 7.25) |
| Georgia | Hispanic | 0.4462 | 0.2976 | 1.53 (0.92; 2.37) |
| Georgia | NH Black | 3.1030 | 1.1129 | 2.81 (2.24; 3.47) |
| Georgia | NH Asian | 0.1537 | 0.0433 | 4.15 (1.29; 10.29) |
| Georgia | NH 2+ races | 0.0164 | 0.0109 | NA |
| Georgia | NH AIAN | <0.0001 | 0.0191 | NA |
| Georgia | NH NHPI | 0.0165 | <0.0001 | NA |
| Hawaii | All races | 1.6325 | 1.2781 | 1.35 (0.62; 2.5) |
| Hawaii | NH White | 0.1002 | 0.0772 | NA |
| Hawaii | Hispanic | 0.1021 | 0.2992 | NA |
| Hawaii | NH Black | 0.1022 | 0.0253 | NA |
| Hawaii | NH Asian | 0.6889 | 0.4764 | 1.77 (0.43; 4.94) |
| Hawaii | NH 2+ races | 0.1030 | 0.1312 | NA |
| Hawaii | NH AIAN | <0.0001 | <0.0001 | NA |
| Hawaii | NH NHPI | 0.5317 | 0.3041 | 2.06 (0.48; 5.65) |
| Idaho | All races | 4.6019 | 1.3284 | 3.58 (2.19; 5.44) |
| Idaho | NH White | 3.5836 | 0.7202 | 5.32 (2.8; 9.22) |
| Idaho | Hispanic | 0.6974 | 0.3418 | 2.33 (0.7; 5.67) |
| Idaho | NH Black | 0.0786 | 0.0475 | NA |
| Idaho | NH Asian | 0.0811 | 0.0941 | NA |
| Idaho | NH 2+ races | 0.0790 | 0.0562 | NA |
| Idaho | NH AIAN | 0.0792 | 0.1736 | NA |
| Idaho | NH NHPI | <0.0001 | <0.0001 | NA |
| Illinois | All races | 7.5330 | 2.2559 | 3.35 (2.94; 3.79) |
| Illinois | NH White | 3.8440 | 0.5567 | 6.96 (5.61; 8.55) |
| Illinois | Hispanic | 1.4946 | 1.0237 | 1.47 (1.14; 1.85) |
| Illinois | NH Black | 1.8446 | 0.5546 | 3.36 (2.56; 4.31) |
| Illinois | NH Asian | 0.3114 | 0.0914 | 3.64 (1.82; 6.6) |
| Illinois | NH 2+ races | 0.0123 | 0.0334 | NA |
| Illinois | NH AIAN | 0.0125 | 0.0072 | NA |
| Illinois | NH NHPI | 0.0125 | <0.0001 | NA |
| Indiana | All races | 6.7011 | 1.5035 | 4.49 (3.65; 5.41) |
| Indiana | NH White | 5.2970 | 0.9022 | 5.93 (4.61; 7.48) |
| Indiana | Hispanic | 0.3514 | 0.1761 | 2.1 (0.97; 3.93) |
| Indiana | NH Black | 0.9492 | 0.3180 | 3.08 (1.86; 4.79) |
| Indiana | NH Asian | 0.0498 | 0.0232 | NA |
| Indiana | NH 2+ races | 0.0208 | 0.0230 | NA |
| Indiana | NH AIAN | 0.0202 | 0.0191 | NA |
| Indiana | NH NHPI | 0.0208 | NA | NA |
| Iowa | All races | 5.9551 | 1.8870 | 3.2 (2.38; 4.19) |
| Iowa | NH White | 5.2292 | 1.2037 | 4.42 (3.14; 6.06) |
| Iowa | Hispanic | 0.2269 | 0.1973 | 1.29 (0.29; 3.36) |
| Iowa | NH Black | 0.2444 | 0.2963 | 0.96 (0.22; 2.55) |
| Iowa | NH Asian | 0.1299 | 0.0378 | 5.67 (0.47; 24.35) |
| Iowa | NH 2+ races | 0.0406 | 0.0395 | NA |
| Iowa | NH AIAN | 0.0411 | 0.0672 | NA |
| Iowa | NH NHPI | 0.0403 | <0.0001 | NA |
| Kansas | All races | 5.3574 | 1.8311 | 2.98 (2.12; 4.08) |
| Kansas | NH White | 3.9778 | 0.8353 | 4.9 (3.16; 7.19) |
| Kansas | Hispanic | 0.6426 | 0.4542 | 1.53 (0.6; 3.09) |
| Kansas | NH Black | 0.4181 | 0.2630 | 1.88 (0.51; 4.79) |
| Kansas | NH Asian | 0.0992 | 0.0749 | NA |
| Kansas | NH 2+ races | 0.0496 | 0.0289 | NA |
| Kansas | NH AIAN | 0.1181 | 0.1605 | NA |
| Kansas | NH NHPI | 0.0484 | <0.0001 | NA |
| Kentucky | All races | 5.0016 | 1.1003 | 4.6 (3.5; 5.98) |
| Kentucky | NH White | 4.1458 | 0.7310 | 5.77 (4.19; 7.8) |
| Kentucky | Hispanic | 0.0906 | 0.1662 | 0.61 (0.08; 1.82) |
| Kentucky | NH Black | 0.7021 | 0.1604 | 4.76 (2.16; 9.3) |
| Kentucky | NH Asian | 0.0323 | 0.0474 | NA |
| Kentucky | NH 2+ races | <0.0001 | <0.0001 | NA |
| Kentucky | NH AIAN | 0.0321 | <0.0001 | NA |
| Kentucky | NH NHPI | <0.0001 | <0.0001 | NA |
| Louisiana | All races | 10.3717 | 3.1707 | 3.29 (2.74; 3.9) |
| Louisiana | NH White | 4.9338 | 0.8740 | 5.74 (4.21; 7.68) |
| Louisiana | Hispanic | 0.2758 | 0.2289 | 1.33 (0.44; 3.01) |
| Louisiana | NH Black | 5.0506 | 2.0830 | 2.44 (1.9; 3.09) |
| Louisiana | NH Asian | 0.0341 | 0.0624 | NA |
| Louisiana | NH 2+ races | 0.0344 | <0.0001 | NA |
| Louisiana | NH AIAN | 0.0342 | 0.0484 | NA |
| Louisiana | NH NHPI | <0.0001 | 0.0145 | NA |
| Maine | All races | 1.0536 | 0.3802 | 3.21 (1.08; 7.5) |
| Maine | NH White | 0.8618 | 0.2295 | 4.61 (1.29; 12.38) |
| Maine | Hispanic | 0.0643 | <0.0001 | NA |
| Maine | NH Black | 0.0646 | 0.1292 | NA |
| Maine | NH Asian | <0.0001 | <0.0001 | NA |
| Maine | NH 2+ races | 0.0655 | <0.0001 | NA |
| Maine | NH AIAN | <0.0001 | <0.0001 | NA |
| Maine | NH NHPI | <0.0001 | <0.0001 | NA |
| Maryland | All races | 6.1901 | 2.0282 | 3.07 (2.48; 3.73) |
| Maryland | NH White | 2.3890 | 0.4003 | 6.15 (3.98; 8.92) |
| Maryland | Hispanic | 0.6074 | 0.6583 | 0.94 (0.54; 1.5) |
| Maryland | NH Black | 2.8841 | 0.8596 | 3.4 (2.48; 4.54) |
| Maryland | NH Asian | 0.2530 | 0.1015 | 2.84 (0.93; 6.75) |
| Maryland | NH 2+ races | 0.0252 | 0.0137 | NA |
| Maryland | NH AIAN | 0.0255 | <0.0001 | NA |
| Maryland | NH NHPI | <0.0001 | <0.0001 | NA |
| Massachusetts | All races | 5.7019 | 1.3515 | 4.25 (3.41; 5.21) |
| Massachusetts | NH White | 4.1981 | 0.6597 | 6.45 (4.87; 8.32) |
| Massachusetts | Hispanic | 0.5993 | 0.3349 | 1.86 (1.02; 3.13) |
| Massachusetts | NH Black | 0.6256 | 0.2577 | 2.54 (1.38; 4.28) |
| Massachusetts | NH Asian | 0.2456 | 0.0671 | 4.23 (1.42; 10.18) |
| Massachusetts | NH 2+ races | 0.0192 | 0.0229 | NA |
| Massachusetts | NH AIAN | 0.0191 | 0.0013 | NA |
| Massachusetts | NH NHPI | <0.0001 | <0.0001 | NA |
| Michigan | All races | 7.2645 | 1.8912 | 3.85 (3.32; 4.44) |
| Michigan | NH White | 4.3668 | 0.8205 | 5.36 (4.35; 6.58) |
| Michigan | Hispanic | 0.2642 | 0.1541 | 1.81 (0.86; 3.3) |
| Michigan | NH Black | 2.4536 | 0.8818 | 2.8 (2.2; 3.52) |
| Michigan | NH Asian | 0.1012 | 0.0219 | 6.36 (1.29; 20.11) |
| Michigan | NH 2+ races | 0.0148 | 0.0134 | NA |
| Michigan | NH AIAN | 0.0466 | 0.0243 | 3.14 (0.31; 13.13) |
| Michigan | NH NHPI | 0.0145 | <0.0001 | NA |
| Minnesota | All races | 3.9854 | 1.0838 | 3.73 (2.75; 4.96) |
| Minnesota | NH White | 3.1238 | 0.4657 | 6.89 (4.67; 9.73) |
| Minnesota | Hispanic | 0.1236 | 0.1773 | 0.81 (0.17; 2.23) |
| Minnesota | NH Black | 0.3780 | 0.2112 | 1.95 (0.77; 3.93) |
| Minnesota | NH Asian | 0.2477 | 0.1146 | 2.38 (0.8; 5.41) |
| Minnesota | NH 2+ races | 0.0225 | 0.0237 | NA |
| Minnesota | NH AIAN | 0.0767 | 0.1133 | 0.92 (0.09; 3.28) |
| Minnesota | NH NHPI | <0.0001 | <0.0001 | NA |
| Mississippi | All races | 11.9856 | 4.1740 | 2.89 (2.33; 3.52) |
| Mississippi | NH White | 5.7330 | 1.1538 | 5.06 (3.59; 6.95) |
| Mississippi | Hispanic | 0.0520 | 0.1714 | NA |
| Mississippi | NH Black | 5.8045 | 2.5602 | 2.29 (1.72; 3.01) |
| Mississippi | NH Asian | 0.1249 | 0.0677 | 2.97 (0.24; 12.88) |
| Mississippi | NH 2+ races | 0.0525 | 0.0147 | NA |
| Mississippi | NH AIAN | 0.2001 | 0.2913 | 0.77 (0.15; 2.14) |
| Mississippi | NH NHPI | <0.0001 | NA | NA |
| Missouri | All races | 6.3405 | 1.4717 | 4.34 (3.51; 5.31) |
| Missouri | NH White | 4.9247 | 0.8761 | 5.68 (4.39; 7.21) |
| Missouri | Hispanic | 0.1739 | 0.1696 | 1.12 (0.35; 2.58) |
| Missouri | NH Black | 1.0788 | 0.3565 | 3.13 (1.88; 4.91) |
| Missouri | NH Asian | 0.0969 | 0.0262 | 5.8 (0.73; 22.46) |
| Missouri | NH 2+ races | 0.0231 | 0.0114 | NA |
| Missouri | NH AIAN | 0.0230 | 0.0047 | NA |
| Missouri | NH NHPI | 0.0227 | 0.0187 | NA |
| Montana | All races | 5.4151 | 2.2380 | 2.49 (1.55; 3.78) |
| Montana | NH White | 3.3617 | 0.8326 | 4.33 (2.16; 7.92) |
| Montana | Hispanic | 0.2435 | 0.2060 | NA |
| Montana | NH Black | 0.1080 | <0.0001 | NA |
| Montana | NH Asian | 0.1105 | 0.0518 | NA |
| Montana | NH 2+ races | 0.1099 | 0.0592 | NA |
| Montana | NH AIAN | 1.4800 | 0.8855 | 1.8 (0.74; 3.56) |
| Montana | NH NHPI | <0.0001 | NA | NA |
| Nebraska | All races | 5.7937 | 2.1483 | 2.76 (1.84; 3.98) |
| Nebraska | NH White | 4.6421 | 1.0610 | 4.55 (2.72; 7.06) |
| Nebraska | Hispanic | 0.6167 | 0.5290 | 1.29 (0.4; 3) |
| Nebraska | NH Black | 0.3033 | 0.3516 | 1.11 (0.17; 3.49) |
| Nebraska | NH Asian | 0.0756 | 0.1172 | NA |
| Nebraska | NH 2+ races | 0.0754 | 0.0348 | NA |
| Nebraska | NH AIAN | 0.0763 | 0.1158 | NA |
| Nebraska | NH NHPI | <0.0001 | <0.0001 | NA |
| Nevada | All races | 7.7300 | 3.1215 | 2.5 (1.95; 3.14) |
| Nevada | NH White | 3.5647 | 0.6374 | 5.8 (3.6; 8.77) |
| Nevada | Hispanic | 1.9570 | 1.6262 | 1.22 (0.8; 1.8) |
| Nevada | NH Black | 0.8411 | 0.3925 | 2.27 (1.06; 4.19) |
| Nevada | NH Asian | 1.1245 | 0.2720 | 4.52 (2.07; 8.95) |
| Nevada | NH 2+ races | 0.0533 | 0.0981 | NA |
| Nevada | NH AIAN | 0.1354 | 0.0671 | 2.92 (0.29; 11.23) |
| Nevada | NH NHPI | 0.0530 | 0.2030 | NA |
| New Hampshire | All races | 1.6809 | 0.6558 | 2.9 (1.11; 6.19) |
| New Hampshire | NH White | 1.4577 | 0.5127 | 3.43 (1.1; 8.31) |
| New Hampshire | Hispanic | 0.0737 | 0.0145 | NA |
| New Hampshire | NH Black | 0.0724 | 0.0539 | NA |
| New Hampshire | NH Asian | 0.0712 | 0.0697 | NA |
| New Hampshire | NH 2+ races | <0.0001 | <0.0001 | NA |
| New Hampshire | NH AIAN | <0.0001 | <0.0001 | NA |
| New Hampshire | NH NHPI | <0.0001 | <0.0001 | NA |
| New Jersey | All races | 12.0527 | 4.4084 | 2.74 (2.44; 3.07) |
| New Jersey | NH White | 5.6872 | 1.2374 | 4.62 (3.78; 5.57) |
| New Jersey | Hispanic | 2.8322 | 1.7287 | 1.65 (1.32; 2.02) |
| New Jersey | NH Black | 2.5457 | 1.0188 | 2.52 (1.97; 3.16) |
| New Jersey | NH Asian | 0.8884 | 0.2510 | 3.63 (2.32; 5.43) |
| New Jersey | NH 2+ races | 0.0598 | 0.0365 | 2.1 (0.3; 6.97) |
| New Jersey | NH AIAN | 0.0175 | <0.0001 | NA |
| New Jersey | NH NHPI | 0.0176 | <0.0001 | NA |
| New Mexico | All races | 6.2166 | 3.5986 | 1.75 (1.28; 2.31) |
| New Mexico | NH White | 1.3238 | 0.3059 | 4.81 (2.04; 9.97) |
| New Mexico | Hispanic | 2.4490 | 1.2936 | 1.95 (1.16; 3.06) |
| New Mexico | NH Black | 0.0662 | 0.1108 | NA |
| New Mexico | NH Asian | 0.0640 | 0.0075 | NA |
| New Mexico | NH 2+ races | 0.0653 | 0.1822 | NA |
| New Mexico | NH AIAN | 2.2448 | 1.9676 | 1.17 (0.71; 1.8) |
| New Mexico | NH NHPI | <0.0001 | <0.0001 | NA |
| New York | All races | 13.2189 | 4.5298 | 2.92 (2.71; 3.15) |
| New York | NH White | 4.9731 | 0.9302 | 5.36 (4.64; 6.16) |
| New York | Hispanic | 3.4806 | 1.9894 | 1.75 (1.52; 2.01) |
| New York | NH Black | 3.5375 | 1.0159 | 3.49 (3.01; 4.02) |
| New York | NH Asian | 1.1809 | 0.5466 | 2.18 (1.69; 2.75) |
| New York | NH 2+ races | 0.0246 | 0.0249 | 1.3 (0.16; 4.49) |
| New York | NH AIAN | 0.0196 | 0.0248 | 1.08 (0.1; 4.06) |
| New York | NH NHPI | 0.0081 | 0.0239 | NA |
| North Carolina | All races | 2.2431 | 0.7184 | 3.15 (2.42; 4) |
| North Carolina | NH White | 1.1151 | 0.2239 | 5.13 (3.35; 7.55) |
| North Carolina | Hispanic | 0.2005 | 0.1724 | 1.21 (0.55; 2.21) |
| North Carolina | NH Black | 0.8430 | 0.2678 | 3.24 (2.06; 4.77) |
| North Carolina | NH Asian | 0.0413 | 0.0149 | 4.29 (0.42; 17.49) |
| North Carolina | NH 2+ races | <0.0001 | <0.0001 | NA |
| North Carolina | NH AIAN | 0.0388 | 0.0078 | NA |
| North Carolina | NH NHPI | <0.0001 | 0.0026 | NA |
| North Dakota | All races | 9.3450 | 3.3986 | 2.84 (1.69; 4.44) |
| North Dakota | NH White | 7.4390 | 1.8535 | 4.27 (2.22; 7.51) |
| North Dakota | Hispanic | 0.2079 | 0.4507 | NA |
| North Dakota | NH Black | 0.2065 | <0.0001 | NA |
| North Dakota | NH Asian | 0.2064 | <0.0001 | NA |
| North Dakota | NH 2+ races | <0.0001 | 0.0282 | NA |
| North Dakota | NH AIAN | 1.2868 | 0.6847 | 2.17 (0.54; 5.58) |
| North Dakota | NH NHPI | <0.0001 | <0.0001 | NA |
| Ohio | All races | 4.8538 | 1.0598 | 4.61 (3.83; 5.48) |
| Ohio | NH White | 3.8523 | 0.7008 | 5.54 (4.47; 6.77) |
| Ohio | Hispanic | 0.0844 | 0.0639 | 1.46 (0.42; 3.51) |
| Ohio | NH Black | 0.8482 | 0.2156 | 4.05 (2.6; 6.01) |
| Ohio | NH Asian | 0.0531 | 0.0216 | 3.12 (0.56; 10.08) |
| Ohio | NH 2+ races | 0.0109 | 0.0255 | NA |
| Ohio | NH AIAN | 0.0113 | 0.0173 | NA |
| Ohio | NH NHPI | <0.0001 | <0.0001 | NA |
| Oklahoma | All races | 6.6928 | 1.6570 | 4.08 (3.11; 5.23) |
| Oklahoma | NH White | 4.4652 | 0.7553 | 6.04 (4.16; 8.45) |
| Oklahoma | Hispanic | 0.4845 | 0.3407 | 1.53 (0.6; 3.17) |
| Oklahoma | NH Black | 0.5024 | 0.1520 | 3.69 (1.35; 8.06) |
| Oklahoma | NH Asian | 0.1043 | 0.0507 | 3.64 (0.24; 16.55) |
| Oklahoma | NH 2+ races | 0.1856 | 0.0273 | NA |
| Oklahoma | NH AIAN | 0.9068 | 0.3236 | 2.95 (1.49; 5.33) |
| Oklahoma | NH NHPI | 0.0401 | 0.1085 | NA |
| Oregon | All races | 1.2744 | 0.6160 | 2.12 (1.31; 3.23) |
| Oregon | NH White | 0.9052 | 0.2151 | 4.46 (2.31; 7.95) |
| Oregon | Hispanic | 0.2088 | 0.2098 | 1.08 (0.32; 2.48) |
| Oregon | NH Black | 0.0291 | 0.0506 | NA |
| Oregon | NH Asian | 0.0744 | 0.0430 | 2.72 (0.2; 11.48) |
| Oregon | NH 2+ races | <0.0001 | 0.0182 | NA |
| Oregon | NH AIAN | 0.0286 | 0.0424 | NA |
| Oregon | NH NHPI | 0.0293 | 0.0396 | NA |
| Pennsylvania | All races | 5.6345 | 1.3990 | 4.04 (3.48; 4.68) |
| Pennsylvania | NH White | 3.8749 | 0.7179 | 5.43 (4.46; 6.57) |
| Pennsylvania | Hispanic | 0.3714 | 0.1849 | 2.07 (1.19; 3.3) |
| Pennsylvania | NH Black | 1.2074 | 0.4081 | 3 (2.19; 3.99) |
| Pennsylvania | NH Asian | 0.1603 | 0.0556 | 3.18 (1.28; 6.63) |
| Pennsylvania | NH 2+ races | 0.0097 | 0.0145 | NA |
| Pennsylvania | NH AIAN | 0.0097 | <0.0001 | NA |
| Pennsylvania | NH NHPI | <0.0001 | <0.0001 | NA |
| Rhode Island | All races | 6.4231 | 1.6755 | 3.97 (2.39; 6.2) |
| Rhode Island | NH White | 4.6780 | 0.7551 | 6.69 (3.34; 12.32) |
| Rhode Island | Hispanic | 0.8788 | 0.1649 | 7.31 (1.41; 23.48) |
| Rhode Island | NH Black | 0.5113 | 0.1751 | 4.45 (0.53; 17.09) |
| Rhode Island | NH Asian | 0.1178 | 0.2059 | NA |
| Rhode Island | NH 2+ races | 0.1157 | 0.0207 | NA |
| Rhode Island | NH AIAN | 0.1153 | <0.0001 | NA |
| Rhode Island | NH NHPI | <0.0001 | <0.0001 | NA |
| South Carolina | All races | 6.1204 | 1.8390 | 3.35 (2.68; 4.15) |
| South Carolina | NH White | 3.2915 | 0.6507 | 5.16 (3.65; 7.11) |
| South Carolina | Hispanic | 0.1321 | 0.1605 | 0.88 (0.24; 2.09) |
| South Carolina | NH Black | 2.6256 | 0.8676 | 3.07 (2.2; 4.14) |
| South Carolina | NH Asian | 0.0266 | 0.0481 | NA |
| South Carolina | NH 2+ races | 0.0261 | <0.0001 | NA |
| South Carolina | NH AIAN | 0.0263 | 0.0064 | NA |
| South Carolina | NH NHPI | <0.0001 | 0.0112 | NA |
| South Dakota | All races | 8.6920 | 3.1946 | 2.81 (1.74; 4.31) |
| South Dakota | NH White | 6.3733 | 1.2726 | 5.31 (2.85; 9.17) |
| South Dakota | Hispanic | 0.1536 | 0.6072 | NA |
| South Dakota | NH Black | 0.1508 | 0.1888 | NA |
| South Dakota | NH Asian | 0.1507 | 0.4051 | NA |
| South Dakota | NH 2+ races | 0.1556 | 0.1591 | NA |
| South Dakota | NH AIAN | 1.6861 | 0.6201 | 3.31 (0.93; 8.45) |
| South Dakota | NH NHPI | <0.0001 | <0.0001 | NA |
| Tennessee | All races | 7.1210 | 1.9507 | 3.67 (3.04; 4.37) |
| Tennessee | NH White | 5.2319 | 0.9670 | 5.46 (4.3; 6.83) |
| Tennessee | Hispanic | 0.2187 | 0.2488 | 0.92 (0.38; 1.83) |
| Tennessee | NH Black | 1.5639 | 0.6233 | 2.55 (1.75; 3.58) |
| Tennessee | NH Asian | 0.0586 | 0.0366 | 2.18 (0.22; 8.19) |
| Tennessee | NH 2+ races | 0.0223 | 0.0113 | NA |
| Tennessee | NH AIAN | 0.0225 | 0.0194 | NA |
| Tennessee | NH NHPI | <0.0001 | 0.0989 | NA |
| Texas | All races | 9.6577 | 2.9475 | 3.28 (3.03; 3.54) |
| Texas | NH White | 3.0860 | 0.5506 | 5.63 (4.8; 6.55) |
| Texas | Hispanic | 5.2701 | 2.0202 | 2.61 (2.36; 2.88) |
| Texas | NH Black | 1.0825 | 0.3152 | 3.46 (2.74; 4.3) |
| Texas | NH Asian | 0.1904 | 0.0389 | 5.15 (2.77; 8.85) |
| Texas | NH 2+ races | 0.0069 | 0.0045 | NA |
| Texas | NH AIAN | 0.0152 | 0.0153 | 1.25 (0.12; 4.37) |
| Texas | NH NHPI | 0.0068 | 0.0171 | NA |
| Utah | All races | 4.2082 | 1.3137 | 3.27 (2.16; 4.74) |
| Utah | NH White | 2.6497 | 0.5537 | 5 (2.81; 8.27) |
| Utah | Hispanic | 0.8718 | 0.4048 | 2.32 (0.96; 4.65) |
| Utah | NH Black | 0.0661 | 0.0095 | NA |
| Utah | NH Asian | 0.1450 | 0.0514 | NA |
| Utah | NH 2+ races | 0.0663 | 0.0355 | NA |
| Utah | NH AIAN | 0.2649 | 0.1145 | 3.13 (0.45; 10.61) |
| Utah | NH NHPI | 0.1463 | 0.0641 | NA |
| Vermont | All races | 0.5108 | 0.3743 | 1.89 (0.18; 7.13) |
| Vermont | NH White | 0.5080 | 0.3148 | 2.52 (0.22; 10.21) |
| Vermont | Hispanic | <0.0001 | <0.0001 | NA |
| Vermont | NH Black | <0.0001 | <0.0001 | NA |
| Vermont | NH Asian | <0.0001 | <0.0001 | NA |
| Vermont | NH 2+ races | <0.0001 | <0.0001 | NA |
| Vermont | NH AIAN | <0.0001 | <0.0001 | NA |
| Vermont | NH NHPI | <0.0001 | <0.0001 | NA |
| Virginia | All races | 3.7909 | 1.0268 | 3.72 (2.94; 4.62) |
| Virginia | NH White | 2.0593 | 0.3368 | 6.25 (4.35; 8.72) |
| Virginia | Hispanic | 0.3352 | 0.2649 | 1.31 (0.66; 2.29) |
| Virginia | NH Black | 1.2154 | 0.3785 | 3.28 (2.18; 4.68) |
| Virginia | NH Asian | 0.1440 | 0.0462 | 3.64 (1.03; 9.27) |
| Virginia | NH 2+ races | 0.0177 | 0.0028 | NA |
| Virginia | NH AIAN | 0.0172 | 0.0058 | NA |
| Virginia | NH NHPI | <0.0001 | <0.0001 | NA |
| Washington | All races | 2.4792 | 0.8638 | 2.91 (2.14; 3.85) |
| Washington | NH White | 1.6196 | 0.2861 | 5.86 (3.72; 8.8) |
| Washington | Hispanic | 0.3977 | 0.2318 | 1.79 (0.9; 3.13) |
| Washington | NH Black | 0.1239 | 0.0910 | 1.66 (0.4; 4.64) |
| Washington | NH Asian | 0.1946 | 0.0813 | 2.8 (0.85; 6.98) |
| Washington | NH 2+ races | 0.0189 | 0.0350 | NA |
| Washington | NH AIAN | 0.0862 | 0.0572 | 1.84 (0.34; 5.34) |
| Washington | NH NHPI | 0.0410 | 0.0554 | NA |
| West Virginia | All races | 3.3386 | 0.7409 | 4.73 (2.72; 7.8) |
| West Virginia | NH White | 3.0653 | 0.6257 | 5.2 (2.87; 8.83) |
| West Virginia | Hispanic | 0.0686 | <0.0001 | NA |
| West Virginia | NH Black | 0.2041 | 0.0708 | NA |
| West Virginia | NH Asian | <0.0001 | 0.0098 | NA |
| West Virginia | NH 2+ races | <0.0001 | <0.0001 | NA |
| West Virginia | NH AIAN | <0.0001 | 0.0623 | NA |
| West Virginia | NH NHPI | <0.0001 | <0.0001 | NA |
| Wisconsin | All races | 5.1974 | 1.3285 | 3.95 (3.08; 4.97) |
| Wisconsin | NH White | 4.0190 | 0.7144 | 5.71 (4.22; 7.55) |
| Wisconsin | Hispanic | 0.4277 | 0.2967 | 1.52 (0.71; 2.81) |
| Wisconsin | NH Black | 0.4677 | 0.1685 | 3 (1.35; 5.82) |
| Wisconsin | NH Asian | 0.1228 | 0.0767 | 1.94 (0.41; 5.65) |
| Wisconsin | NH 2+ races | 0.0243 | <0.0001 | NA |
| Wisconsin | NH AIAN | 0.1138 | 0.0733 | 2.24 (0.33; 7.82) |
| Wisconsin | NH NHPI | 0.0247 | 0.0282 | NA |
| Wyoming | All races | 3.4128 | 1.8758 | 1.97 (0.82; 3.95) |
| Wyoming | NH White | 2.4350 | 0.5757 | 5.06 (1.53; 13.07) |
| Wyoming | Hispanic | 0.2188 | 0.3201 | NA |
| Wyoming | NH Black | <0.0001 | <0.0001 | NA |
| Wyoming | NH Asian | 0.2214 | 0.1999 | NA |
| Wyoming | NH 2+ races | <0.0001 | 0.0248 | NA |
| Wyoming | NH AIAN | 0.5303 | 0.7580 | NA |
| Wyoming | NH NHPI | <0.0001 | <0.0001 | NA |
