## Supplementary material for "Fairness and efficiency considerations in COVID-19 vaccine allocation strategies: a case study comparing front-line workers and 65-74 year olds in the United States": 2022-09-12-Vaccine_prioritization_Supplementary_Table7_with95CI.docx

**S7 Table. Relative number of years of life saved comparing a policy allocating vaccines to 65-74 year olds (policy S) to one allocating vaccines to front-line workers (policy F). Results shown for varying values of *R*, the increased risk of infection for front-line workers.** The ratios for an R value of 1·56 correspond to those shown in Table 4. "NH" stands for "Non-Hispanic", "2+ races" stands for "Two or more races", "Native" stands for "American Indian or Alaska Native", "NHPI" stands for "Native Hawaiian or Other Pacific Islander".

| Race/ethnicity | **Ratio Y (95% uncertainty interval)** | | | | |
| --- | --- | --- | --- | --- | --- |
|  | ***R* = 1** | ***R* = 1·56** | ***R* = 2** | ***R* = 3** | ***R* = 4** |
| All races | 4·9 (4·77; 5·04) | 3·14 (3·06; 3·23) | 2·45 (2·39; 2·52) | 1·63 (1·59; 1·68) | 1·23 (1·19; 1·26) |
| Non-Hispanic White | 8·49 (8·13; 8·85) | 5·44 (5·21; 5·67) | 4·24 (4·07; 4·43) | 2·83 (2·71; 2·95) | 2·12 (2·03; 2·21) |
| Hispanic | 2·89 (2·75; 3·04) | 1·85 (1·76; 1·95) | 1·45 (1·38; 1·52) | **0·96** (**0·92**; 1·01) | **0·72** (**0·69**; **0·76**) |
| Non-Hispanic Black | 4·32 (4·08; 4·57) | 2·77 (2·62; 2·93) | 2·16 (2·05; 2·29) | 1·44 (1·36; 1·53) | 1·08 (1·02; 1·14) |
| Non-Hispanic Asian | 4·35 (3·82; 4·93) | 2·79 (2·45; 3·16) | 2·17 (1·91; 2·45) | 1·45 (1·28; 1·64) | 1·09 (**0·96**; 1·23) |
| Non-Hispanic 2+ races | 2·01 (1·36; 2·81) | 1·29 (**0·88**; 1·82) | 1·00 (**0·68**; 1·4) | **0·67** (**0·45**; **0·94**) | **0·50** (**0·34**; **0·70**) |
| Non-Hispanic Native | 2·27 (1·86; 2·74) | 1·45 (1·19; 1·75) | 1·13 (**0·93**; 1·37) | **0·76** (**0·62**; **0·92**) | **0·57** (**0·47**; **0·68**) |
| Non-Hispanic NHPI | 1·14 (**0·74**; 1·66) | **0·73** (**0·47**; 1·07) | **0·57** (**0·37**; **0·83**) | **0·38** (**0·25**; **0·55**) | **0·28** (**0·18**; **0·41**) |
