## Supplementary figures and images for "Fairness and efficiency considerations in COVID-19 vaccine allocation strategies: a case study comparing front-line workers and 65-74 year olds in the United States"

### SupplFigure1.tif

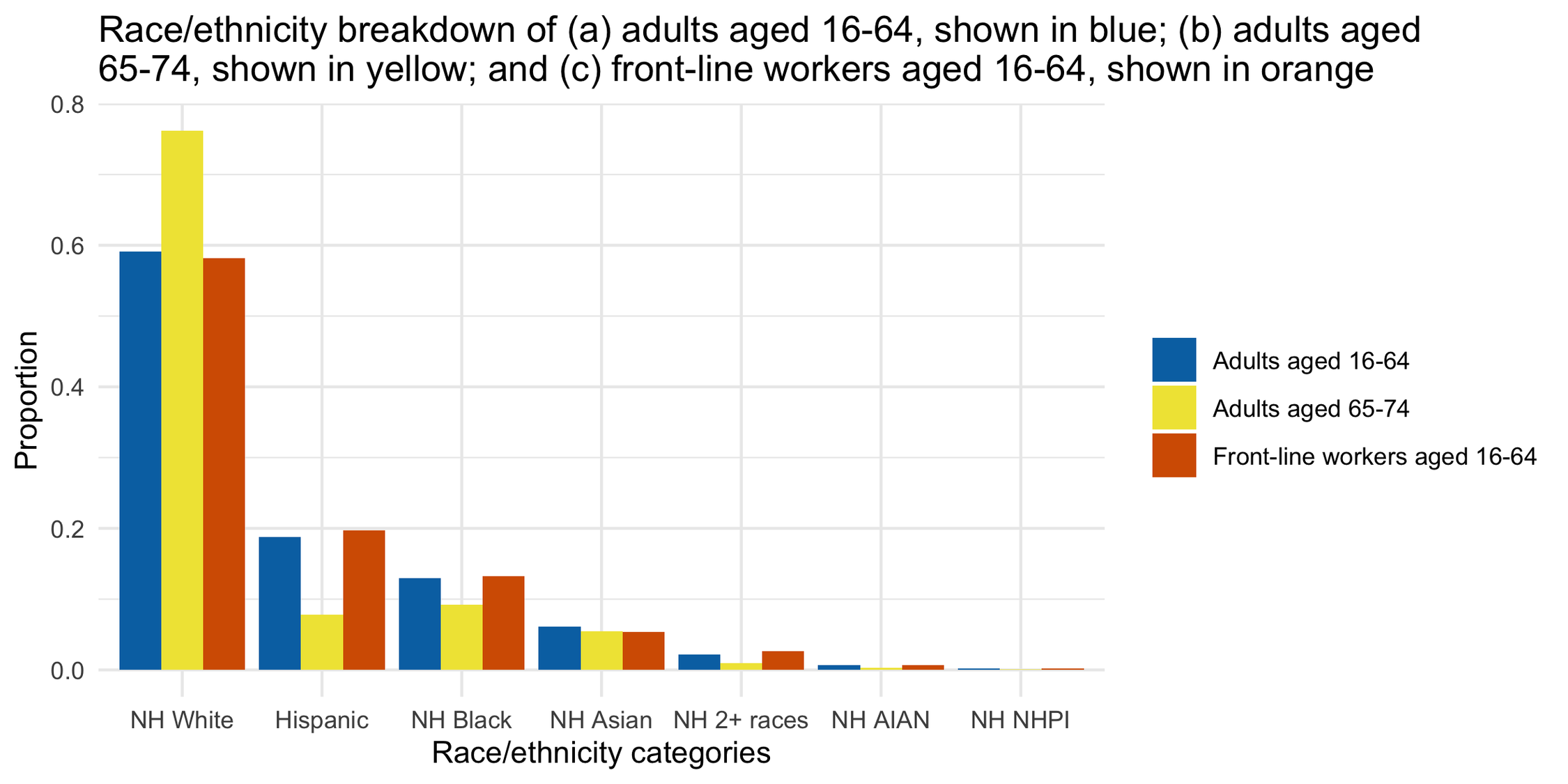

### SupplFigure2.tif

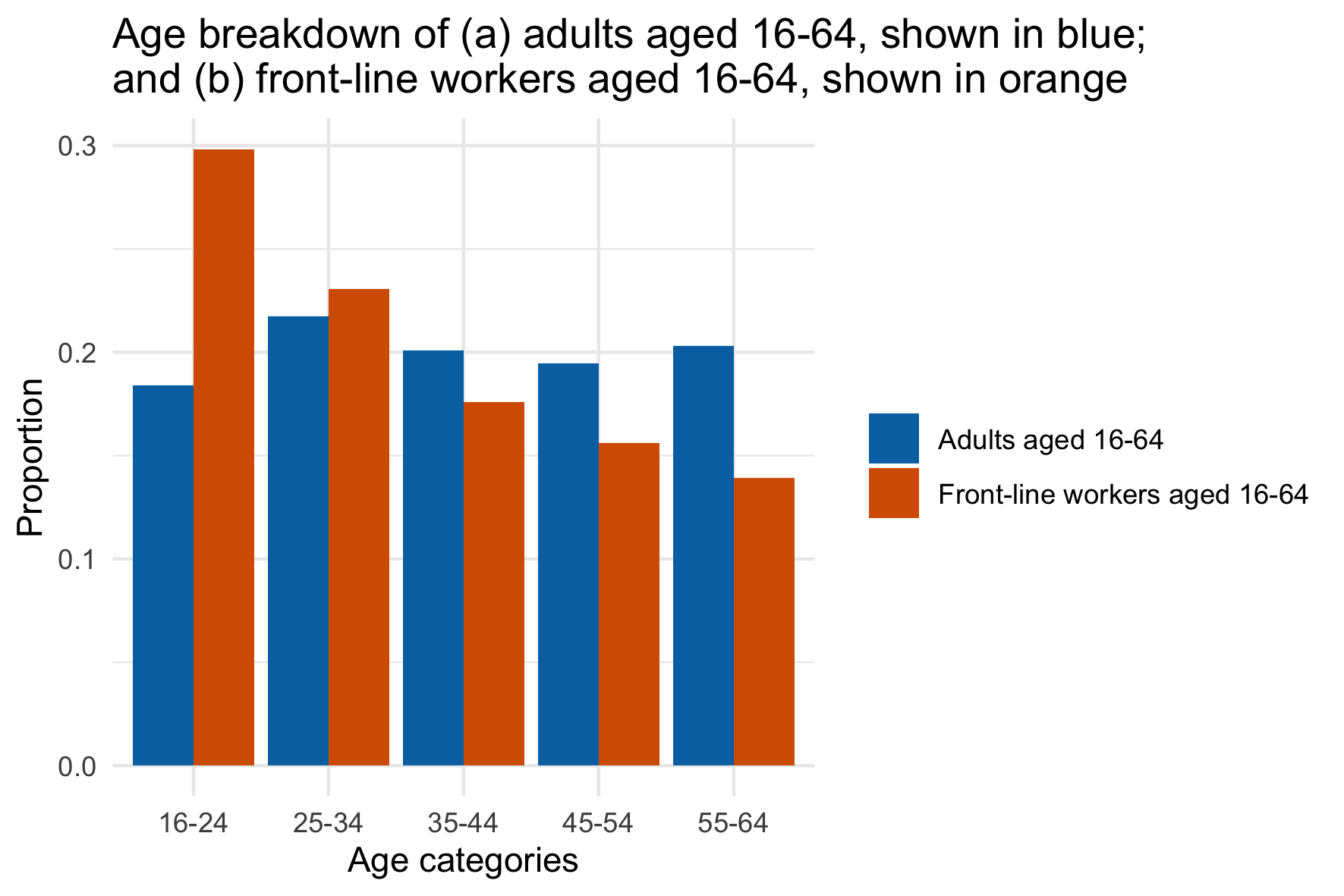

### SupplFigure3.tif

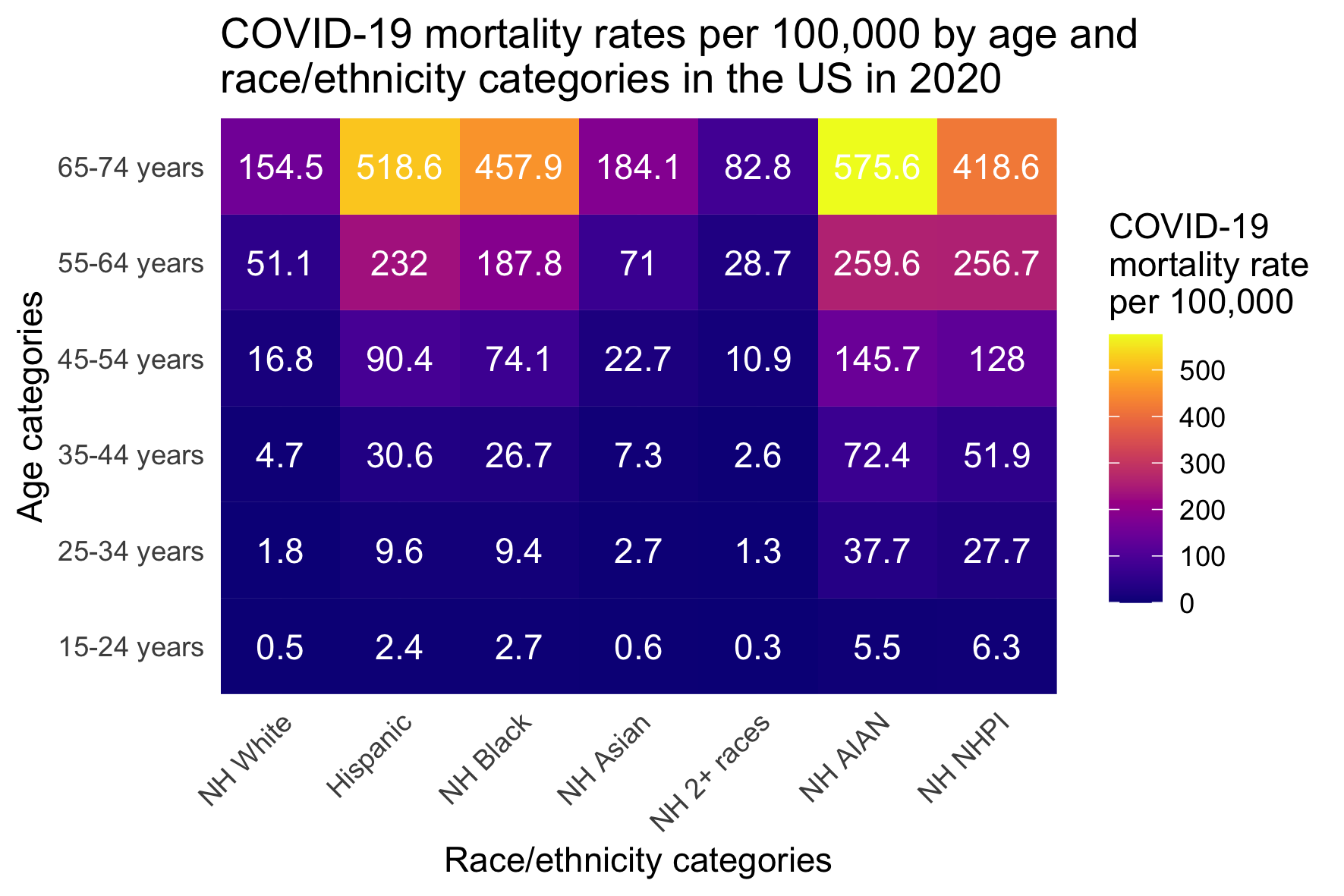
